# CSF proteome profiling reveals highly specific biomarkers for dementia with Lewy bodies

**DOI:** 10.1101/2023.07.10.23292447

**Authors:** Marta del Campo, Lisa Vermunt, Carel FW Peeters, Yanaika S. Hok-A-Hin, Alberto Lleó, Daniel Alcolea, Mirrelijn van Nee, Sebastiaan Engelborghs, Anne Sieben, Alice Chen-Plotkin, David J Irwin, Wiesje M van der Flier, Afina W Lemstra, Charlotte E Teunissen

## Abstract

Diagnosis of dementia with Lewy bodies (DLB) is challenging and biofluid biomarkers specific for DLB are highly needed. Here we use proximity extension-based multiplex assays to establish the specific cerebrospinal fluid (CSF) proteomic changes that underlie DLB in an unprecedented well-characterized cohort of 109 DLB patients, 235 patients with Alzheimeŕs disease (AD) and 190 controls. We identified more than 50 CSF proteins dysregulated in DLB, which were especially related to myelination processes. An enzyme involved in dopamine biosynthesis (L-amino acid decarboxylase, DDC) was the strongest dysregulated protein in DLB (>1.5 fold-change vs.CON or AD; q<1E-16) and could discriminate DLB from controls and AD patients with high accuracy (AUC: 0.91 and 0.81 respectively). We modelled a CSF protein panel containing only seven of these markers, which discriminate DLB from AD with higher performance (AUC: 0.93, 95%CI: 0.86-0.98). We developed custom multiplex assays for six of these markers (DDC, CRH, MMP-3, ABL1, MMP-10 and THOP1); and validated their performance in independent cohorts (n=329; AUCs: 0.68-0.90), including an autopsy cohort (n=76; AUCs: 0.90-0.95). This extensive and unique DLB CSF proteome study depicts specific protein changes underlying DLB pathophysiology. It translates these findings into a custom CSF biomarker panel able to identify DLB patients with high accuracy in different independent cohorts, providing new testing opportunities for diagnostic settings and clinical trials.

## 1. Introduction

Dementia with Lewy Bodies (DLB) is one of the most common forms of dementia in the aged population after Alzheimer’s disease (AD)^1^ and is clinically characterized by cognitive fluctuations, visual hallucinations, parkinsonism and rapid eye movement sleep behavior disorder. DLB is pathologically characterized by the intraneuronal accumulation of α-synuclein (α-syn) in Lewy bodies in the neocortex^2^. The clinical and pathological presentation strongly overlap with AD, challenging differential diagnosis and leading to a large proportion of miss- or undiagnosed DLB patients ^3–5^.

Limited number of biomarkers have been widely analyzed to date in DLB. Despite previous studies on α-syn in cerebrospinal fluid (CSF) showing conflicting results^6–8^, recent developments using real-time quaking-induced conversion (RT-QuIC) assays allow to detect α-syn brain proteinopathy in CSF and skin ^9–12^. These novel assays can discriminate DLB from control or AD patients with high accuracy^9–12^. However, α-syn pathology is not unique for DLB patients and more than 40% of AD cases can present with this comorbid pathology^13^. Similarly, previous studies have shown that the core CSF biomarkers used to support AD diagnosis (amyloid β peptide (Aβ_1-42_), total tau (tTau) and phosphorylated tau (pTau))^14^ provide limited diagnostic accuracy for discriminating DLB from AD since they are also abnormal in almost 25-40% of DLB patients due to the presence of comorbid AD pathology ^15–18^. Additional markers reflecting different, specific and unique aspects of DLB pathophysiology are needed, which could be useful for different contexts of use in both clinical settings (e.g., prognosis, differential diagnosis, disease monitoring) and trials (e.g., patient selection, stratification, treatment efficacy).

CSF proteome profiling allows to identify changes covering a wide range of biological processes *in vivo*. As observed within the AD field, such analysis can open new insights into the molecular mechanisms involved in disease pathogenesis and reveal promising biomarker candidates^19–21^. The few DLB proteomic studies performed to date did not yield many biomarker candidates, which could be due to the limited sample size (30-40 samples per group) relative to the DLB heterogeneity^22–25^. We here employed a high-throughput proteomics method (immune-based proximity extension assay (PEA)) that allows analysis of large cohorts, with the additional advantage that custom multiplex immunoassays including the markers of interest can be smoothly developed for large scale validation^22, 27^. We have applied this workflow to (i) define novel CSF proteomic changes underlying DLB pathogenesis and (ii) to identify, develop and validate multiplex biomarker assays that could aid in the specific diagnosis of DLB.

## 2. Methods

### 2.1 Participants

An overview of the study design is presented in figure 1. The total discovery cohort (n=534) included CSF samples from patients diagnosed with DLB (n=109), AD (n=235) and 190 cognitively unimpaired controls (CON; table 1). Most of the samples were selected from the Amsterdam Dementia Cohort (ADC) and DEvELOP^26, 27^. To enrich for DLB dementia cases, additional CSF samples from the Center for Neurodegenerative Disease Research at the University of Pennsylvania were included (49 DLB and 18 AD)^28^. Three additional independent CSF cohorts were used for validation of the customized panels (see methods below): clinical validation cohort 1 (from the ADC: 54 DLB, 55 AD, and 55 controls)^26^, clinical validation cohort 2 (from the Sant Pau Initiative on Neurodegeneration (SPIN) cohort: 55 DLB, 55 AD, and 55 controls)^29^ and an autopsy confirmed cohort (from BIODEM, UANtwerp and the neurobiobank of the Institute Born-Bunge (IIB) / UAntwerp: 17 DLB and 30 AD)^29, 30^. An additional 29 cognitively unimpaired controls from BIODEM-UAntwerp were included in this cohort but these were not autopsy confirmed^30, 31^. CSF was collected by lumbar puncture and processed and stored at all sites in agreement with the JPND-BIOMARKAPD guidelines^32^.

**Figure 1.**
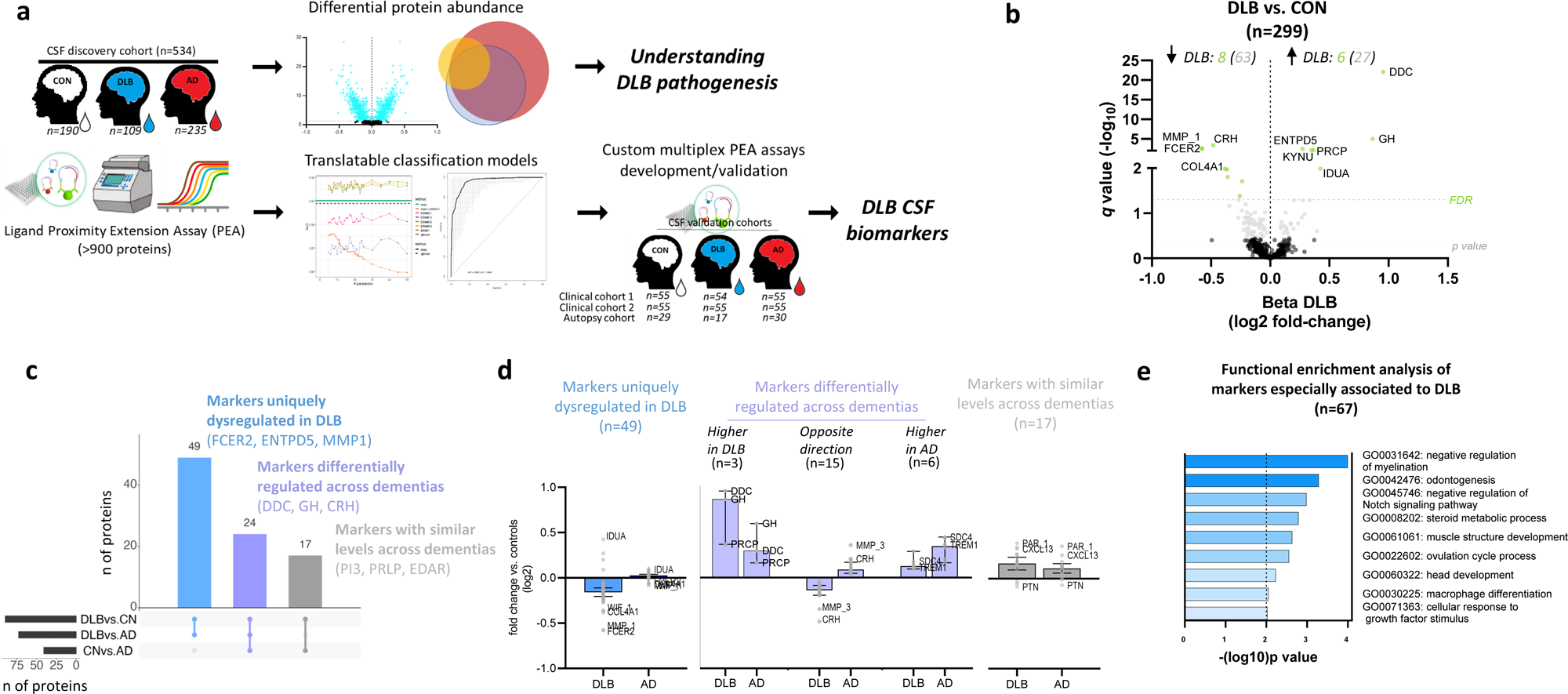
Study overview and differential abundance of CSF proteins in DLB. a, Protein levels in CSF from cognitively unimpaired controls (white), DLB (blue) and AD (red) were measured by antibody-based PEA technology. Differential CSF protein abundance as well as classification models were investigated. Custom multiplex PEA assays containing the markers identified within the classification panels were developed and validated in three independent validation cohorts. b, Volcano plot shows the CSF proteins that are differentially regulated in DLB vs. controls. Each dot represents a protein. The beta coefficients (log2 fold-change) are plotted versus q values (-log10-transformed). Proteins significantly dysregulated after adjusting for false discovery rate (FDR, q < 0.05) are coloured in light green and those with nominal significance (p < 0.05) are coloured in grey. The name of the top 10 significant dysregulated CSF proteins and the top 5 with the strongest effect sizes are annotated. The total number of proteins that are down-regulated (left) or up-regulated (right) is indicated. Horizontal dotted line indicates the significance threshold. c, UpSet plot indicate which of the proteins dysregulated between DLB and controls are also dysregulated between DLB and AD or AD and controls. d, Bar plots depict the direction of changes of the different proteins identified when compared to controls within the groups depicted by the UpSet plot. e, Bar graphs depicting the biological pathways enriched in those protein dysregulated in DLB. Dot line represent the significant threshold (p <0.01). The corresponding GO number and biological process is defined in the right side. Stronger colours represent higher significant enrichment. CON, cognitively unimpaired controls; DLB, Dementia with Lewy bodies, AD, Alzheimer’s disease.

**Table 1.** Demographic characteristics.

|  | Discovery cohort |  |  | Validation cohort 1 |  |  | Validation cohort 2 |  |  | Autopsy cohort |  |  |
| --- | --- | --- | --- | --- | --- | --- | --- | --- | --- | --- | --- | --- |
|  | CON | DLB | AD | CON | DLB | AD | CON | DLB | AD | CON | DLB | AD |
|  | (n=190) | (n=109) | (n=235) | (n=55) | (n=54) | (n=55) | (n=55) | (n=55) | (n=55) | (n=29) | (n=17) | (n=30) |
| Age, years (Mean, SD) | 58 (8)<sup>a,b</sup> | 69 (8)<sup>b,c</sup> | 66 (8)<sup>a,c</sup> | 58 (4)<sup>a,b</sup> | 69 (5)<sup>b,c</sup> | 66 (8)<sup>a,c</sup> | 62 (7)<sup>a,b</sup> | 76 (5)<sup>b,c</sup> | 72 (6)<sup>a,c</sup> | 63 (5)<sup>a,b</sup> | 76 (7)<sup>c</sup> | 71 (11)<sup>c</sup> |
| Sex (M, %) | 120 (63%) | 91 (83%) | 139 (59%) | 33 (60%) | 45 (83%) | 32 (58%) | 23 (41%) | 33 (60%) | 21 (38%) | 16(55%) | 14 (82%) | 15 (50%) |
| MMSE (Mean, SD)<sup>1</sup> | 28 (2)<sup>a,b</sup> | 22 (6)<sup>b,c</sup> | 21 (5)<sup>a,c</sup> | 28 (1)<sup>a,b</sup> | 23(4)<sup>b,c</sup> | 20 (3)<sup>a,c</sup> | 29(1)<sup>a,b</sup> | 22(5)<sup>c</sup> | 22(4)<sup>c</sup> | 30(1)<sup>b</sup> | 17(7) | 14(8)<sup>c</sup> |
| APOE4 (+/n, %)<sup>2</sup> | 47/186 (25%) | 52/97 (54%) | 134/226 (59%) | 15/55 (27%) | 28/51 (55%) | 39/55 (71%) | 14/55 (25%) | 18/54 (33%) | 24/54 (44%) | na | na | na |
| CSF Aβ42, pg/mL<sup>3</sup> | 1121 (218)<sup>a,b</sup> | 764 (372)<sup>b,c</sup> | 603 (123)<sup>a,c</sup> | 1283 (301) / 1629 (372)<sup>a,b</sup> | 851 (528) / 751 (338)<sup>b,c</sup> | 566 (154) / 494 (159)<sup>a,c</sup> | 1194 (503)<sup>a,b</sup> | 672 (454)<sup>b,c</sup> | 537 (206)<sup>a,c</sup> | 1108 (284)<sup>a,b</sup> | 461(398)<sup>c</sup> | 404 (206)<sup>c</sup> |
| CSF tTau, pg/mL<sup>3</sup> | 211 (95)<sup>b</sup> | 297 (193)<sup>c</sup> | 746 (431)<sup>a,c</sup> | 292 (244) / 182 (91)<sup>b</sup> | 293 (165) / 214 (108)<sup>c</sup> | 460 (352) / 341 (115)<sup>a,c</sup> | 257 (125)<sup>a,b</sup> | 432 (486)<sup>b,c</sup> | 740 (439)<sup>a,c</sup> | 203 (91)<sup>a,b</sup> | 368 (314)<sup>c</sup> | 535 (420)<sup>c</sup> |
| CSF pTau, pg/mL<sup>3</sup> | 38 (15)<sup>a,b</sup> | 52 (28)<sup>b,c</sup> | 92 (36)<sup>a,c</sup> | 54 (28) / 15 (8)<sup>b</sup> | 48 (23) / 16 (12)<sup>b,c</sup> | 63 (44) / 36(22)<sup>a,c</sup> | 38 (21)<sup>a,b</sup> | 63 (80)<sup>b,c</sup> | 117 (84)<sup>a,c</sup> | 47 (19)<sup>b</sup> | 57 (28) | 72(38)<sup>c</sup> |

Data are median (interquartile range) unless otherwise specified.
<sup>1</sup>MMSE score was used as a measure of cognitive function it was missing for 12, 2, 6 and 41 participants in the discovery, validation 1, 2 and autopsy cohorts respectively. Note that there was MMSE from only 2 control cases in the autopsy cohort
<sup>2</sup>APOE status was missing for 25, 3 and 2 participants in the discovery, validation 1 and 2 cohorts respectively.
<sup>3</sup>Biomarker data coming from Luminex analysis in the discovery cohort was transformed using appropriate Passing-Babock transformation formulas (see methods). In validation cohort 1 the first values correspond to data obtained with Innotech and the second values correspond to data obtained with Elecsy. CSF Biomarker data from validation cohort 2 was obtained with Lumipulse.
*p* < 0.05 vs.DLB<sup>a</sup>, vs. AD<sup>b</sup>, or vs. CON<sup>c</sup>.
DLB, Dementia with Lewy Bodies; AD, Alzheimer’s disease; SD, Standard desviation;M, Male.

All participants of every cohort underwent standard neurological and cognitive assessments and diagnosis was assigned according to international consensus criteria for DLB^33, 34^ and AD^35^. The neuropathological validation cohort included cases with a definite diagnosis according to international neuropathological examination guidelines for DLB^33,34^ and AD^36^. Mini-Mental State Examination (MMSE) was used as a measure of global cognition. Levels of CSF Aβ_42_, tTau and pTau(181) (‘AD CSF biomarkers’) were used to support AD diagnoses. These markers were analyzed locally as part of the diagnostic procedure using commercially available kits (VUmc and UAntwerp: ELISA INNOTEST Aβ(1-42), hTAUAg, phospho-Tau(181P, Fujirebio, Ghent, Belgium) or VUmc: Aβ(1-42), t-TAUAg, phospho-Tau181 Elecsys biomarker assays (Roche Diagnostics GmbH); Penn: Luminex xMAP INNO-BIA AlzBio3; Luminex Corp, Austin, TX; SPIN: Lumipulse G600, Fujirebio)^37–39^. Positive CSF AD biomarker profile was defined locally as increased tTau/Aβ_42_ ratio in the cohorts from ADC (>0.46) and Pennsylvania (>0.30); and low Aβ_42/40_ ratio (<0.062) and high total tau (>456pg/ml) or p-tau (>63pg/ml) in the SPIN cohort ^29, 40–42^.

In the discovery cohort, DLB neuropathology was confirmed in 14 DLB patients (13%) by autopsy (supplementary table 1). From those DLB patients that did not have autopsy confirmation (n=95), clinical diagnosis was supported by FPCIT single-photon emission computed tomography (DAT scan) in 23 patients (24% of the total DLB patients with clinical diagnosis, supplementary table 1). Autopsy data or any supporting biomarker information was not available for 48 DLB patients (44%, supplementary table 1). In the DLB group, 72 patients (66%) had a negative CSF AD biomarker profile, 34 patients (32%) had a positive CSF AD profile, and three patients (3%) did not have both AD CSF biomarkers available. Of note, four out of 13 of the DLB patients with autopsy confirmation (31%) had also a positive AD CSF profile. From all the DLB patients, only five DLB patients had medication for parkinsonian symptoms, 55 did not have such a treatment and this information was not available for 49 cases.

Diagnosis of AD patients was supported by a positive AD CSF biomarker profile in 233 patients (99%) and in nine patients this was confirmed by autopsy (which include the 1% of patients that either had a negative AD CSF biomarker profile or did not have CSF biomarkers available). The control group included individuals with subjective cognitive decline, in whom objective cognitive and laboratory investigations were normal (i.e., criteria for MCI, dementia, or any other neurological or psychiatric disorder not fulfilled) with additionally negative AD CSF biomarkers in all cases^26, 43–45^.

In the clinical validation cohorts 1 and 2, DAT scans supported DLB diagnoses in 19 and 21 patients respectively (35% and 38% of the total DLB patients, supplementary table 1). DAT scans were not available for 18 and 27 DLB patients from validation cohort 1 and 2 respectively (33% and 49%, supplementary table 1). 55% and 52% of the DLB patients from validation cohorts 1 and 2 respectively had a negative AD CSF biomarker profile. In validation cohort 1, 49 AD patients (89%) had a positive AD CSF profile but, unlike the discovery cohort, 19 AD patients (35%) were CSF pTAU negative. In validation cohort 2, 50 AD patients (91%) had a positive AD CSF profile, 2 patients had negative AD CSF biomarker profile, and no CSF information was available for 3 AD patients. All control cases except two from the validation cohort 1 had negative AD CSF profile.

The neuropathological validation cohort included cases with a definite diagnosis according to international neuropathological examination guidelines for DLB ^33, 34^ and AD^36^. Within the DLB group coexisting AD pathological changes (n=7) or cerebrovascular lesions (n=3) were reported in 10 cases. Coexisting cerebrovascular lesions (n=8), TDP pathology (n=1) or cerebral amyloid angiopathy (n=1) were reported within the AD neuropathological group. The control group of this cohort was not autopsy confirmed. It consisted of volunteers, mainly spouses of patients who visited the memory clinic. The inclusion criteria for these volunteers was: (1) no neurological or psychiatric antecedents; (2) no organic disease involving the central nervous system following extensive clinical examination; and (3) normal neuropsychological exam. Exclusion criteria consisted of brain tumors, large cerebral infarction/bleeding, strategic infarctions, other neurodegenerative diseases, severe head trauma, epilepsy, brain infections, severe depression, unregulated diabetes mellitus, untreated thyroid disorders, or any severe somatic comorbidity that interferes with study participation^46^. Only one case within the control group had positive CSF Aβ_42_ value, but its CSF pTau and tTau levels were normal.

Patient demographics and clinical and biochemical values from all cohorts used in this study are listed in table 1. The studies were approved by the Institutional Ethical Review Boards of each center. Informed consent was obtained from all subjects or their authorized representatives.

### 2.2 CSF protein profiling

As part of our large-scale discovery project^21^, CSF proteins (979) were quantified using the 11 specific and validated multiplex antibody-based protein panels based on the proximity extension assay (PEA) that were available at the time in which the analysis was performed as previously described (Cardiometabolic, Cardiovascular II and III, cell regulation, development, immune response, inflammation, metabolism, neurology, oncology II and organ damage; Olink Proteomics, Uppsala, Sweden; supplementary table 2)^47^. Each panel contains reagents to measure up to 92 unique proteins, though 30 proteins can be measured in several panels (replicates). Briefly, samples were randomized across plates containing appropriate intra- and inter-plate quality controls (QC) from the manufacturer and measured in two different rounds. Each round included 16 bridging samples covering different clinical groups, which were used for reference sample normalization to control for potential batch effects. Each assay has an experimentally determined lower limit of detection (LOD) estimated as three standard deviations above noise level from the negative controls that are included on every plate. Only proteins with values over the lower limit of detection (LOD) in at least 85% of the samples were selected for further statistical analysis, in which remaining raw values under LOD (2.4% of all measurements) were kept as provided by manufacturer. A total of 665 proteins (642 unique proteins) were ultimately included for statistical analysis of the discovery cohort (supplementary table 2).

### 2.3 Development of custom PEA assays

Custom designed multiplex-PEA assays were developed by the manufacturer following standardized protocols^48–50^. We developed assays to measure six out of the seven proteins selected upon the classification analysis described in section 2.4. Besides the corresponding clinical samples, each plate included four CSF QC samples, a negative control and three calibrators used for normalization. QC samples and calibrators were measured in triplicate. Each custom assay has an experimentally determined LOD defined as for the discovery panels. Precision (intra- and inter-assay CV) were calculated using the 4 CSF QC samples. No cross-reactivity between assays for specific proteins was detected. Assay parameters including LOD, detectability and CVs are included in supplementary table 3. Samples from validation cohorts were randomized across plates and normalized for any plate effects using the built-in inter-plate controls according to manufacturers’ recommendations. Protein abundance was reported in NPX values.

### 2.4 Statistical analysis

All data preprocessing and analyses were conducted using R version 3.5.3 and SPSS version 25. Between-group analyses for the demographic variables were performed using two-sided one-way analysis of variance in normally distributed continuous data or Pearson’s chi-square test for categorical variables. Analysis of covariance was performed when an association between classical AD CSF biomarker and age and/or sex were detected. Adjustment for multiple testing was performed using Bonferroni method. Non-Gaussian distributed data were analyzed using Kruskal-Wallis Test. For the CSF proteome data, differences in protein abundance between pairs of clinical groups were evaluated using nested linear models as previously described, in which for each individual protein feature, we assessed if its addition to a base model containing age and gender contributed to model fit^21, 51^. For each pairwise comparison, multiplicity was taken into account by controlling the False Discovery Rate (FDR)^52^ at *q* ≤ 0.05 based on the number of features analyzed. We next evaluated which CSF protein combination (CSF panels) could best discriminate the groups of interest while keeping the number of markers to the minimum, so that they can be ultimately translated into small, practical custom panel^21^. For this purpose, binary classification signatures (DLB vs. CON and DLB vs. AD) were constructed by way of penalized generalized linear modeling (GLM) with an elastic net penalty (a linear combination of lasso and ridge penalties) in the discovery CSF cohort using the glmnet package and including age and sex as covariates^21, 51, 53^. This penalty enables estimation in settings where the feature to sample ratio is too high for standard generalized linear regression. Moreover, it performs automatic feature decorrelation as well as feature-selection. For each classification exercise, we compare multiple models which reflect (a) a grid of values for the elastic-net mixing parameter, reflecting strong decorrelation to a pure logistic lasso regression and (b) a grid of values reflecting the maximum number of proteins that may be selected under each model (21 markers maximum). The former grid (a) considers that we have little information on the collinearity burden in the data. The latter grid (b) considers that we want to keep the number of selected proteins relatively low for the future development of customized panels. The optimal penalty parameters in the penalized models were determined based on (balanced) 10-fold cross-validation of the model likelihood^21, 51^. The cross-validation was performed with balanced folds, by which each fold has an outcome group ratio close to the corresponding ratio in the full data set, also referred to as stratified cross-validation. Predictive performance of all models was assessed by way of (the comparison of) Receiver Operating Characteristic (ROC) curves and Area Under the ROC Curves (AUCs). The model with the highest AUC and lowest number of markers for each classification signature was selected. The fold-based selection proportions for each marker were assessed to identify and select the most promising markers within each model (i.e., features that are stably selected across each individual fold thereby minimizing potential overfitting). To reflect the manual selection pressure for these final marker sets, each final logistic signature was subjected to a ridge-regularization with a penalty parameter of 0.1. The performance (AUC) was evaluated by internal validation: repeated 5-fold cross-validation with 1000 repeats. The 95% confidence interval around the resulting AUCs was based on resampling quantiles (percentile method). External validations assessed the performance of the final models with the markers of interest in the validation cohorts using ROC analysis.

Non-parametric correlation analysis was performed to understand the associations between the proteins within the CSF panels and the classical AD CSF biomarkers or cognitive function (MMSE score) using the complete discovery cohort without stratifying per diagnostic category and conditioning on age and sex as covariates. Innotest values generated for the Amsterdam Dementia Cohort were adjusted for drift over time as previously described^54^. UPENN values had lower means for Aβ_42_ on the Innotest, which were first linearly transformed to normalize to the same mean. Passing-Bablock transformation formulas were calculated based on individuals with both Luminex and Innotest values for Aβ_42_ (n=32), tatou (n=32) and pTau (n=27) and used the formulas to estimate the equivalent Innotest values for those samples measured with Luminex platform only (transformed_ Aβ_42_ = (Luminex_ Aβ42*4.65) - 36.23; transformed_tTau = (Luminex_tTau*5.28) - 2.03; transformed_ptau = (Luminex_pTau*1.88) + 27.36).

Functional enrichment analysis was performed using Metascape^52^ selecting GO Biological Processes as ontology source. All the CSF proteins optimally analyzed with Olink arrays (n=645 protein gene products) were included as the enrichment background. Default parameters were used for the analysis in which terms with a *p*-value < 0.01, a minimum count of 3, and an enrichment factor > 1.5 were collected and grouped into clusters based on their membership similarities.

## 3. Results

An overview of the study design is presented in figure 1. We included a total of 534 participants in the discovery cohort, a subset of patients previously analyzed in our previous CSF proteomic study^21^. Custom multiplex panels were developed and validated in two independent clinical cohorts (validation cohort 1: n=164; validation cohort 2: n=165) and one autopsy-confirmed cohort (n=76). The demographic characteristics and AD CSF biomarkers are described in table 1. Cognitively unimpaired controls were younger in all the cohorts analyzed. Cases included in validation cohort 2 and the autopsy cohort were overall older than the other cohorts.

### 3.1 CSF proteins differentially regulated DLB compared to controls and AD

CSF proteome profiling revealed a total of 14 proteins differentially regulated in DLB compared to controls after correcting for multiple testing (Figure 1b, Extended data table 1 (ED Table 1), q <0,05). Six of these proteins were upregulated in DLB (DDC, GH, IDUA, PRCP, KYNU and ENTPD5) and eight proteins were downregulated (CRH, FCER2, MMP1, COL4A1, WIF1, PAM, VEGFA and CTSC, Figure 1b, ED Table 1). Of note, up to 90 proteins showed nominal significant differences between DLB and controls (*p* < 0.05; 27 upregulated and 63 downregulated in DLB Figure 1b, ED Table 1). Three of these proteins had replicates measured across different panels within the proteomic platform (see methods), which highly correlated with each other (r > 0.6; supplementary fig. 1). The top 5 differentially regulated proteins (median *q*: 4.37^-04^) are involved in the biosynthesis of dopamine and serotonin (DDC, or so called AADC), growth control (GH), corticotropin release from pituitary gland (CRH), immune function (FCER2) and extracellular remodeling (MMP1). DDC showed the strongest effects (β = 0.95; fold-change: 1.9; *q* = 8.08^-23^) followed by GH, MMP1, FCER2 and CRH (fold-changes >1.5; figure 1b and ED table 1).

To understand whether the protein changes identified were specific for DLB, we next analyzed if the proteins showing nominal differences in DLB were also differentially changed between DLB and AD as well as between AD and controls (ED table 1). UpSet plot indicate that up to 49 proteins (55%) were uniquely dysregulated in DLB (e.g., FCER2, ENTPD5, MMP1, Figure 1c), which were mostly downregulated (figure 1d). Up to 17 proteins (19%) did not differ between dementias (e.g., PI3, PRLP, EDAR), which likely represent general dementia markers. Interestingly, 24 proteins (27%) were changed between DLB and AD, but also between AD and controls (e.g., DDC, GH, CRH). When compared to controls, we observed that most of these markers showed opposite direction of changes in DLB and AD (e.g., CRH, MMP3), three markers were more strongly associated to DLB (e.g., DDC, GH, PRCP) and six markers showed stronger associations to AD (e.g., SCD4, TREM1, Figure 1d). Functional enrichment analysis showed that the markers especially associated to DLB pathophysiology (i.e., those that were specifically changed in DLB as well as those that showed opposite direction or higher differences compared to the AD group, n=67) were reflecting different biological processes including myelination regulation, tooth development, Notch signaling, steroid metabolism, muscle structure development or ovulation cycle (Figure 1e).

### 3.2 CSF protein markers discriminate DLB from cognitively unimpaired controls and AD dementias

DDC, the strongest dysregulated marker, could discriminate DLB from controls with high accuracy (AUC 0.91, 95% CI: 0.88-0.94, figure 2a). DDC could also discriminate DLB from AD with good but lower performance (AUC 0.81, 95% CI: 0.76-0.86; figure 2a). To investigate whether a minimal combination of biomarkers could discriminate DLB from AD patients with higher accuracies, we next performed classification analysis, followed by internal cross-validation (CSF panels, figure 1a). We identified a panel of 7 CSF proteins including DDC that discriminated DLB from controls and AD with higher accuracies than DDC alone (DLB vs CN AUC: 0.95, 95% CI: 0.89-0.99, DeLonǵs test p<0.001; DLB vs AD: AUC: 0.93, 95% CI: 0.86-0.98, DeLonǵs test p<0.0001, figure 2b,c). The model contained proteins that were dysregulated in DLB compared to both controls and AD (DDC, FCER2, CRH), as mentioned above, as well as one with nominal significant differences (MMP-3; figure2c). The model also included proteins that were not changed in DLB but were specifically upregulated in AD (ABL1, MMP-10 and THOP1; figure 2c), as previously reported in our PEA-AD CSF study^21^. It is worth noting that similar accuracies were obtained when DLB patients with a positive or negative AD CSF biomarker profile (based on tau/Aβ42 ratio) were analyzed separately, indicating that AD pathology comorbidities did not influence the performance of the model (supplementary figure 2). Sensitivity analysis including only those patients that were not under any parkinsonian medication (n=55) showed similar results (supplementary figure 3), suggesting that the changes observed were not driven by levodopa associated treatment. When compared to the CSF biomarkers used to support AD diagnosis (i.e., Aβ_42_/tTau), we observed that the performance of the CSF panel could better discriminate DLB from controls and showed similar AUCs for the discrimination of DLB from AD (Figure 2d). We performed correlation analysis in the complete discovery cohort to understand how these markers relate to cognitive function or classical AD biomarkers (Figure 2e). For the subset of markers especially dysregulated in DLB, we observed that DDC and MMP-3 correlated with MMSE, albeit weakly. Moderate correlations were observed between CSF (p)Tau levels and CRH and MMP-3. Weak negative correlations were detected between Aβ_42_ and DDC levels. As expected, the strongest correlations with AD CSF biomarkers and MMSE were observed for those markers that were specifically dysregulated in AD (ABL1, MMP10 and THOP1).

**Figure 2.**
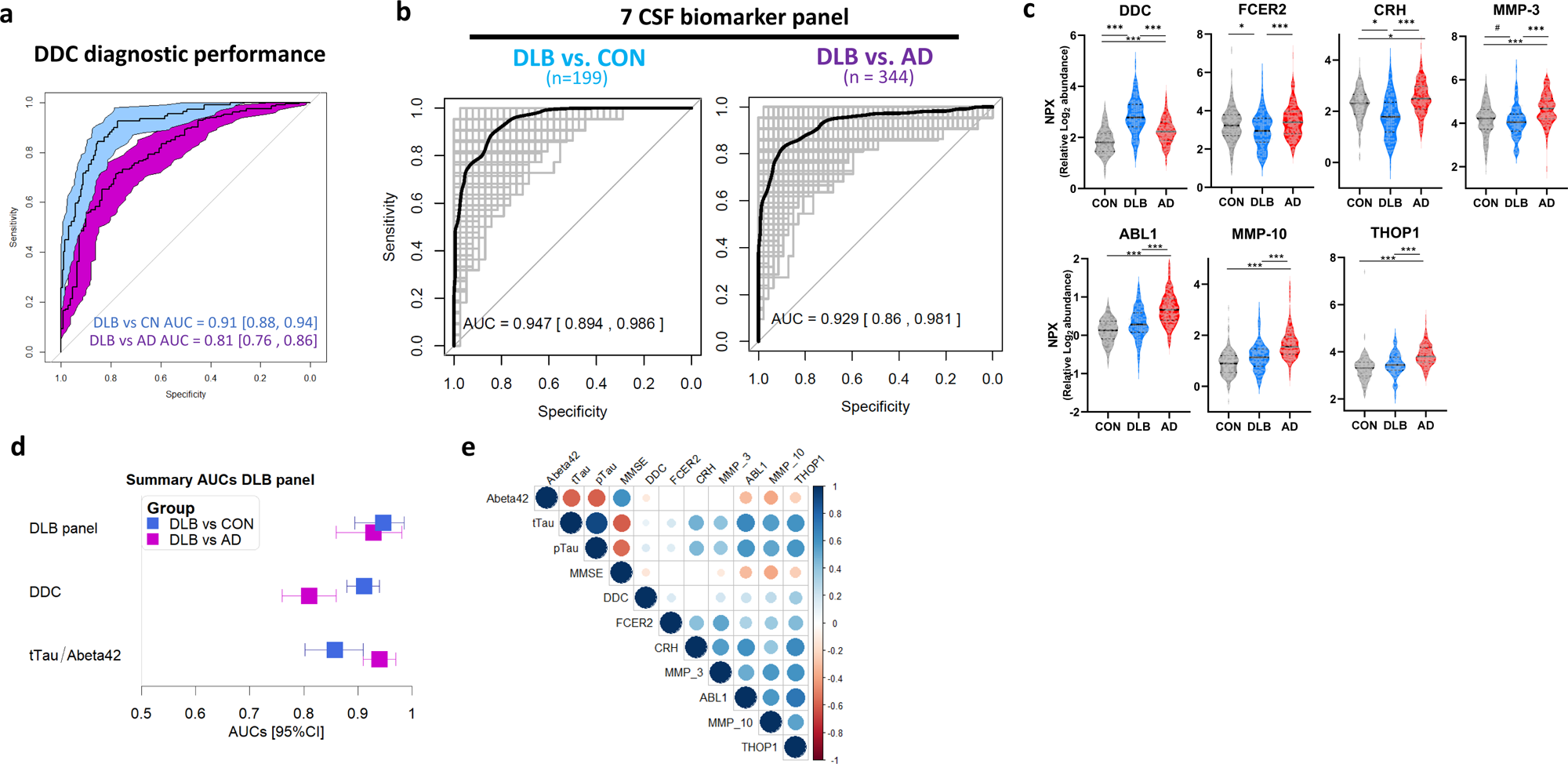
CSF biomarker panels for specific diagnosis of DLB. a) Receiver operating characteristic (ROC) curves depict the performance of CSF DDC discriminating DLB (n = 109) from controls ((n = 190, blue) and AD (n=235, purple). b) ROC curves depicting the performance of 7 CSF biomarker panel discriminating DLB from controls and AD. Black line is the mean area under the curve (AUC) over all re-samplings (1000 repeats of 5-fold cross-validation, grey lines). Inserts outline corresponding AUC and 95% CI. c) Violins represent the abundance (log2 NPX) of the different CSF proteins within the DLB biomarker panel. Horizontal black and dash lines indicate median and interquartile range of the protein abundance. d) Forest plot depicts the different AUC and 95%CI obtained with the CSF DLB biomarker panel, CSF DDC or CSF tTau/Abeta42 biomarkers in the comparison between DLB and controls (blue) or AD (purple). e) Correlation matrix heatmap representing the Spearman’s correlation coefficient in-between the proteins selected in each panel, the classical AD CSF biomarkers and ratios and MMSE score. Significant associations are depicted by circles. *q<0.05, **q<0.01, ***q<0.001. DLB, dementia with Lewy Bodies; AD, Alzheimer’s disease; CON, cognitively unimpaired controls.

The proteins involved in the 7 CSF biomarker panel are related to different pathways including dopamine biosynthesis (DDC), immune function (FCER-2), intra and extracellular remodeling (MMP-3 and MMP-10, ABL1), regulation of the hypothalamic–pituitary–adrenal axis (CRH) and neuropeptide degradation (THOP1).

### 3.3 Development of custom multiplex PEA assays and validation of CSF protein panels

To validate the performance of our discovery findings, we developed custom multiplex PEA-panels measuring six out of the seven proteins from the DLB diagnostic panel, including DDC. Custom assays showed low coefficients of variation (mean intra- and inter-assay CVs of 5% and 9% respectively) and >90% detectability (supplementary table 3). We next analyzed three independent CSF cohorts using these custom assays. We observed that the protein fold changes between DLB and controls or AD dementia obtained in the three validation cohorts correlated highly with those obtained in the discovery cohort (r > 0.70, figure 3a). Of note, the effect size of DDC change in the clinical validation cohort 2 was lower to the ones obtained with the discovery and clinical validation cohort 1 and the autopsy cohort (figure 3a). In the three cohorts, the DLB CSF panel showed slightly higher accuracies than DDC alone in discriminating DLB from controls and AD (Figure 3b,c). The performance of both DDC and the panel in the clinical validation cohort 1 were similar to those observed in the discovery phase (AUCs > 0.86; Figure 3b,c). In the second validation cohort, the accuracies were lower compared to those obtained in the discovery and the other validation cohorts, especially when discriminating DLB from AD (AUC*_DDC_* DLBvs.CON: 0.81; AUC*_DDC_* DLBvs.AD: 0.59; AUC*_custom panel_* DLBvs.CN: 0.90; AUC*_custom panel_* DLBvs.AD: 0.68; Figure 3b,c). The accuracies of this second clinical validation cohort were not improved when only DLB cases with abnormal DAT scan were analyzed (Supplementary figure 4). Analysis of the autopsy cohort with these multiplex custom assays reported similar high accuracies as those of the discovery and clinical validation cohort 1 (AUC*_DDC_* DLBvs.CON: 0.95; AUC*_DDC_* DLBvs.AD: 0.86; AUC*_custom panel_* DLBvs.CN: 1.00; AUC*_custom panel_* DLBvs.AD: 0.90; Figure 3b,c).

**Figure 3.**
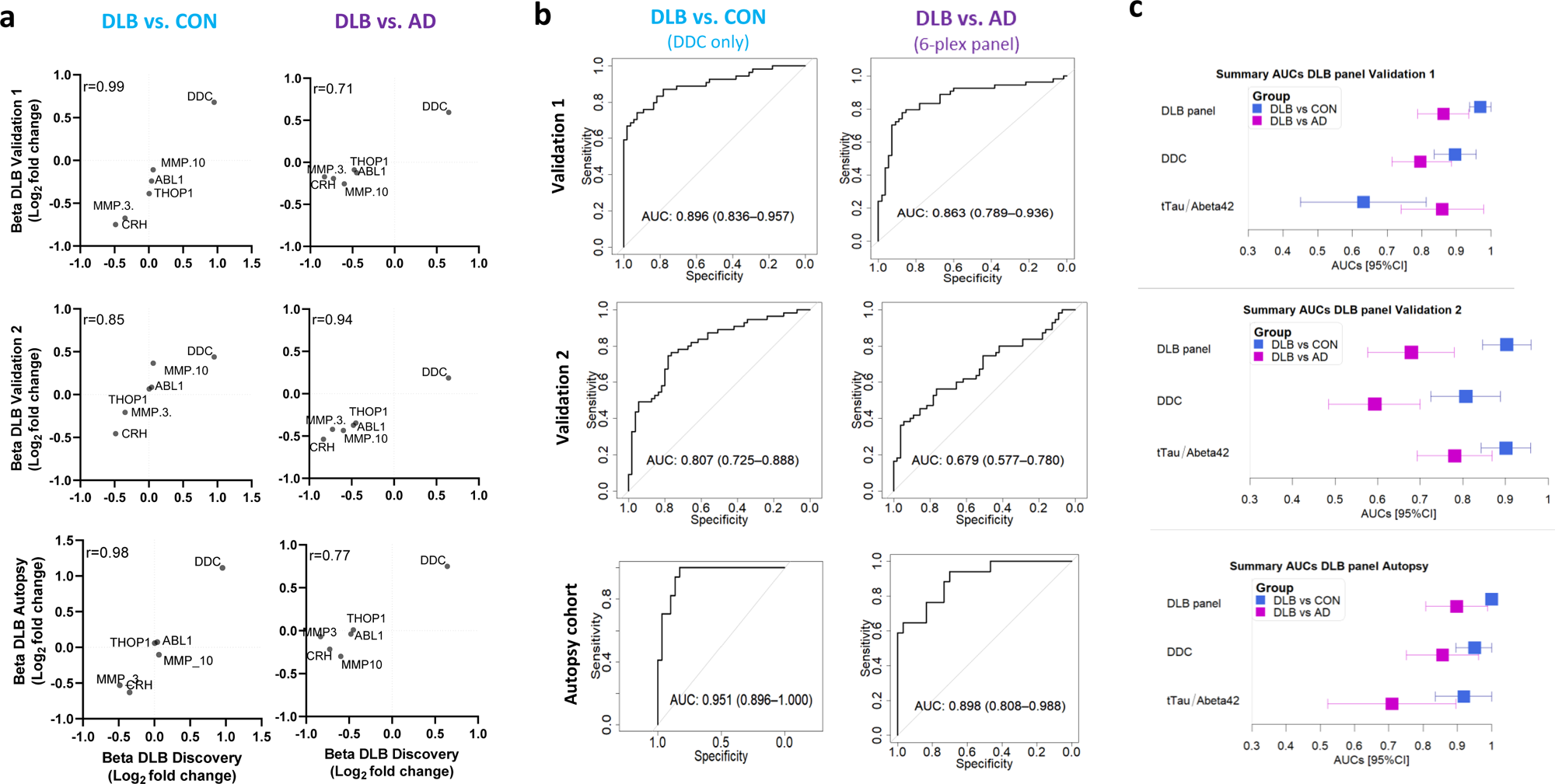
Development and validation of custom CSF biomarker panels for DLB diagnosis in independent cohorts. a) Scatter plots show the correlation between the beta-coefficients obtained in the discovery phase to those obtained with the custom assays in the clinical validation cohorts 1 and 2 and the autopsy confirmed cohort. Insert indicate the spearman correlation coefficient. b) Receiver operating characteristic (ROC) curves depicting the performance of DDC or the CSF biomarker panel discriminating DLB from controls or AD respectively using the custom assays across the different clinical and autopsy validation cohorts. Inserts outline corresponding AUC and 95% CI. Forest plots depict the different AUC and 95%CI obtained with the CSF DLB biomarker panel, CSF DDC or the CSF tTau/Abeta42 ratio in the comparison between DLB and controls (blue) or AD (purple). DLB, dementia with Lewy Bodies; AD, Alzheimer’s disease; CON, cognitively unimpaired controls.

## 4. Discussion

The extensive and unique DLB CSF proteome profiling performed in this study revealed several novel proteins specifically changed in DLB. We translated these findings into a six CSF protein custom panel that discriminated DLB patients from cognitively unimpaired controls and AD with high accuracies (AUCs ≥ 0.90), which was validated in independent external cohorts, including one with neuropathology confirmation. The proteins identified reflect known biological processes associated to DLB pathophysiology such as the biosynthesis of dopamine.

Biofluid-based biomarkers specifically associated to DLB are strongly needed not only to improve timely diagnosis and diagnostic accuracy, but also to monitor the different biological mechanisms involved in DLB pathophysiology and as surrogate markers for clinical trials^55^. To the best of our knowledge this is the largest DLB CSF proteome study performed to date (>100 samples per group)^23, 56^, in which we included not only DLB and cognitively unimpaired controls, but also samples from patients with AD dementia. We detected up to 90 proteins dysregulated in the CSF of DLB cases, but only 14 survived correction for multiple testing. Larger sample sizes might still be required to detect additional biomarker candidates due to clinical heterogeneity in DLB. Thanks to the inclusion of the AD dementia group, we could show that up to 55% of these 90 proteins were dysregulated in DLB specifically (e.g., FCER2, MMP1, WIF1). We identified an additional subset of CSF proteins (27%) that were differentially regulated in both DLB and AD patients compared to controls, but with different protein abundance also between DLB and AD. While some of these were decreased in DLB and increased in AD (e.g., CRH and MMP3), some were especially dysregulated in AD as previously reported (e.g., SDC4, TREM1)^21^, and some were more prominently dysregulated in DLB (e.g., DDC, GH). These last shared but different protein profiles might be explained by the clinicopathological overlap across these two dementias^13, 57^. Most of the proteins identified were downregulated in DLB compared to controls, a proteomic pattern observed in a previous proteome study^56^ but also in recent DLB transcriptomic studies^58–61^. It has been suggested that such strong downregulated pattern in DLB could be due to demyelination processes^60^. In line with those findings, we observed that the proteins dysregulated in DLB were especially enriched in processes associated to a negative regulation of myelination. Furthermore, previous research indicates that alpha-synuclein can induce myelin loss in neurons and oligodendroglia (precursor) cells^62, 63^. Importantly, α-synuclein-induced myelination deficits are involved in the development of multiple system atrophy^63^. The importance of this mechanism for DLB remains to be investigated further.

The strongest dysregulated CSF protein in the DLB group was DDC, an enzyme involved in the biosynthesis of dopamine, which lends biological support to our biomarker discovery design^64^. Previous studies have shown that serum DDC enzyme activity is elevated in patients using levodopa with peripheral decarboxylase inhibitors (PDI), underpinning the effect of dopaminergic treatment on DDC activity^65^. It is important to note that the increase of CSF DDC levels detected in this study are likely not driven by levodopa/PDI treatment as i) PDIs do not cross the blood-brain barrier and can thus not influence DDC activity/levels in the brain and ii) we obtained similar results when only DLB patients that did not have any parkinsonism medication were included in the analysis (n=55). Dysfunction of the dopaminergic system because of nigrostriatal degeneration is a well-established pathophysiological feature of DLB^66^. Previous studies have shown that nigrostriatal degeneration as well as antagonist of dopamine receptors increase DDC mRNA and activity in different models^64^. This data does not only align with our findings, but also suggests that the increase DDC levels might be a response to the nigrostriatal degeneration (and subsequent loss of dopaminergic receptors) and could thus be a very relevant biomarker for DLB diagnosis and disease monitoring. We could validate the high discriminatory accuracy for DLB vs controls (AUC: 0.91) in three independent cohorts, including a neuropathological one (AUCs 0.81-0.95). Considering that current DLB diagnostic guidelines include supportive imaging biomarkers as proxy of nigrostriatal degeneration^4^, it would be of interest to specifically analyze whether CSF DDC measurements could be an alternative or complementary diagnostic test to classical imaging scans. DDC measurements have the additional advantage of being quantitative over the binary classification with the RT-QuIC assay, meaning that, DDC measurements might be relevant to track disease staging and for monitoring treatment responses.

To translate the CSF proteome findings into practical biomarker tools for routine diagnostics and clinical trials, we applied classification analysis and identified a panel of seven CSF proteins to discriminate DLB from controls and AD dementia with high accuracy (AUC of 0.95 and 0.93 respectively). This panel combined proteins associated to DLB (e.g., DDC, CRH, MMP3, FCER2) but also proteins specifically related to AD (ABL1, MMP-10, THOP1)^67–72^, likely explaining the higher performance to discriminate these two dementias compared to DDC alone. To validate these findings in independent cohorts, we successfully developed custom multiplex assays for six out of the seven selected markers. The protein effect sizes obtained with these custom assays in the three validation cohorts correlated well with those obtained in the discovery cohort (r coefficients ranging between 0.70 and 0.99), and the high discriminative values were mostly validated (AUCs ≥ 0.80), supporting the relevance and robustness of our findings. In the clinical validation cohort 2 we observed, however, lower accuracy when discriminating DLB vs. AD (AUC 0.68). The heterogeneity of the clinical diagnosis of DLB may explain the differences observed across these cohorts. Understanding which factors may influence CSF DDC levels is of paramount importance for the potential future implementation of this marker and the corresponding DLB diagnostic panel.

Despite the unprecedented number of samples and proteins analyzed in this DLB specific study, there are still relevant limitations. Considering the clinicopathological overlap with AD^13, 57^, we cannot exclude that potential misdiagnosis of DLB patients may have influenced our discovery results. However, samples were collected in well-characterized biobanks from specialized memory units, and DLB diagnosis was either autopsy confirmed or supported by DAT scans in more than one third of the patients. Moreover, our sensitivity analysis showed similar diagnostic accuracies in DLB cases with positive AD CSF biomarker profile, and we further validated the biomarker panels in a CSF autopsy confirmed cohort. It would be of interest to analyze the performance of these markers in other α-synucleinopathies (e.g., Parkinsońs disease), other dementia types with motor dysfunctions (e.g., progressive supranuclear palsy, corticobasal degeneration) or other conditions with dopamine deficiency. Future studies are still needed to define the clinicopathological correlations between the biomarker panel and different measures associated with DLB pathophysiology, including other relevant markers, such as α-syn^9, 11, 12, 73, 74^. This will help to define their potential context of use within different settings (prognosis, diagnosis, monitoring, etc.). We envision that CSF DDC and the panel developed within this study could be relevant complementary diagnostic tests reflecting different biological aspects associated to DLB pathophysiology. This unique quality may make them suitable to improve diagnosis and staging along the DLB continuum but also to monitor treatment response in clinical trials targeting different mechanisms^75, 76^.

Overall, we identified CSF biomarkers specifically associated with DLB by a unique and extensive CSF proteome profiling, opening new insights into the pathophysiology of this dementia. The protein panels discriminate DLB from controls and AD dementia with high accuracy, which we have translated into custom assays and validated in independent cohorts, including one with autopsy confirmation. These biomarkers and panels are ready to be employed to define their added value and potential context-of-use in clinical settings and trials within the context of DLB. The use of an antibody-based technology allowed us to overcome the cross-technology gap often encountered in biomarker studies^77^ and efficiently translate our discovery findings into customized assays. Current studies are ongoing to validate the CSF biomarkers using alternative immunoanalytical platforms. The workflow employed in this study may ultimately facilitate bench-to-bedside translation of biofluid based biomarker findings and could thus be also relevant for other research fields.

## Data Availability

The data that support the findings of this study are available from the authors on reasonable request.

## Acknowledgments

The authors acknowledge Prof. Dr. Jean-Jacques Martin (Institute Born-Bunge, Antwerp) for the neuropathological characterization of the Antwerp cohort.

## 6. Key words

Dementia with Lewy bodies, Alzheimer’s disease, differential diagnosis, cerebrospinal fluid, biomarkers, proteins.

## 7. Data availability

The data that support the findings of this study are available from the authors on reasonable request. The studies were approved by the Institutional Ethical Review Boards of each center (Discovery cohort: VUmc: AD CSF biobank METC number 00-211; University of Pennsylvania: language and cognitive impairment in parkinson’s disease and parkinson’s disease with dementia or dementia with lewy bodies IRB069801; or under the Parelsnoer initiative 2009-170.)

