## Supplementary material for "CSF proteome profiling reveals highly specific biomarkers for dementia with Lewy bodies": ED Table 1

**Extended data Table 1. CSF Proteins differentially regulated across the different diagnostic groups.**

| | | PROTEINS DIFFERENTIALY REGULATED IN THE DISCOVERY COHORT | | | | | | | | |
|---|---|---|---|---|---|---|---|---|---|---|
| | | DLB vs CON (n=299) | | | DLB vs AD (n=344) | | | AD vs CON (n=425) | | |
| Uniprot | Name | Effect | *p*- value | *q*- value | Effect | *p*- value | *q*- value | Effect | *p*- value | *q*- value |
| P20711 | DDC | 0.954 | 1.22E-25 | **8.08E-23** | 0.643 | 2.05E-19 | **4.55E-17** | 0.297 | 3.10E-07 | **5.03E-06** |
| P01241 | GH | 0.864 | 2.99E-08 | **9.95E-06** | 0.292 | 2.01E-02 | **2.91E-02** | 0.597 | 1.86E-07 | **3.18E-06** |
| P06850 | CRH | -0.484 | 1.97E-06 | **4.37E-04** | -0.833 | 2.82E-22 | **1.09E-19** | 0.208 | 2.71E-03 | **1.11E-02** |
| P06734 | FCER2 | -0.575 | 1.56E-05 | **2.59E-03** | -0.677 | 7.31E-11 | **1.33E-09** | -0.012 | 9.00E-01 | 9.23E-01 |
| O75356 | ENTPD5 | 0.271 | 2.80E-05 | **3.11E-03** | 0.216 | 1.73E-05 | **4.88E-05** | 0.056 | 1.88E-01 | 2.82E-01 |
| P03956 | MMP-1 | -0.579 | 2.71E-05 | **3.11E-03** | -0.430 | 3.84E-04 | **8.18E-04** | -0.082 | 4.32E-01 | 5.33E-01 |
| P42785 | PRCP | 0.367 | 7.40E-05 | **6.99E-03** | 0.139 | 2.27E-02 | **3.23E-02** | 0.162 | 1.12E-02 | **3.41E-02** |
| Q16719 | KYNU | 0.347 | 8.41E-05 | **6.99E-03** | 0.132 | 6.42E-02 | 8.17E-02 | 0.129 | 3.53E-02 | 7.77E-02 |
| P02462 | COL4A1 | -0.384 | 1.55E-04 | **1.03E-02** | -0.379 | 7.60E-06 | **2.33E-05** | -0.065 | 3.41E-01 | 4.44E-01 |
| P35475 | IDUA | 0.423 | 1.53E-04 | **1.03E-02** | 0.224 | 8.43E-03 | **1.32E-02** | 0.094 | 2.27E-01 | 3.23E-01 |
| Q9Y5W5 | WIF-1 | -0.366 | 1.77E-04 | **1.07E-02** | -0.457 | 1.66E-08 | **1.08E-07** | -0.044 | 5.26E-01 | 6.15E-01 |
| P19021 | PAM | -0.360 | 2.84E-04 | **1.57E-02** | -0.527 | 5.63E-12 | **1.78E-10** | 0.060 | 3.39E-01 | 4.42E-01 |
| P15692 | VEGFA | -0.239 | 3.84E-04 | **1.96E-02** | -0.359 | 8.36E-11 | **1.43E-09** | 0.003 | 9.42E-01 | 9.54E-01 |
| P53634 | CTSC | -0.257 | 8.67E-04 | **4.12E-02** | -0.186 | 2.47E-03 | **4.19E-03** | -0.055 | 3.23E-01 | 4.25E-01 |
| P12644 | BMP-4 | -0.269 | 1.15E-03 | 5.09E-02 | -0.313 | 2.80E-06 | **9.81E-06** | 0.003 | 9.66E-01 | 9.74E-01 |
| O43927 | CXCL13 | 0.333 | 1.35E-03 | 5.15E-02 | -0.091 | 3.57E-01 | 3.98E-01 | 0.260 | 6.34E-04 | **3.65E-03** |
| P19957 | PI3 | 0.274 | 1.39E-03 | 5.15E-02 | 0.155 | 5.10E-02 | 6.64E-02 | 0.115 | 8.33E-02 | 1.50E-01 |
| P10144 | GZMB | 0.201 | 1.29E-03 | 5.15E-02 | 0.165 | 6.00E-04 | **1.19E-03** | 0.005 | 9.12E-01 | 9.33E-01 |
| Q96D42 | KIM1 | -0.208 | 1.81E-03 | 6.35E-02 | -0.322 | 4.04E-08 | **2.30E-07** | 0.023 | 6.25E-01 | 7.01E-01 |
| P08254 | MMP-3 | -0.346 | 2.08E-03 | 6.92E-02 | -0.726 | 2.17E-17 | **2.89E-15** | 0.360 | 5.68E-06 | **6.19E-05** |
| P55285 | CDH6 | -0.239 | 2.77E-03 | 8.76E-02 | -0.410 | 1.57E-10 | **2.55E-09** | 0.066 | 2.34E-01 | 3.31E-01 |
| Q9NQX5 | NPDC1 | -0.045 | 3.16E-03 | 9.55E-02 | -0.042 | 5.52E-04 | **1.11E-03** | 0.006 | 5.85E-01 | 6.65E-01 |
| P51888 | PRELP | 0.131 | 3.61E-03 | 9.64E-02 | 0.042 | 2.16E-01 | 2.50E-01 | 0.056 | 7.99E-02 | 1.46E-01 |
| Q9BQT9 | CLSTN3 | -0.197 | 3.35E-03 | 9.64E-02 | -0.346 | 4.38E-11 | **1.00E-09** | 0.061 | 1.58E-01 | 2.47E-01 |
| O00451 | GFRA2 | -0.130 | 3.62E-03 | 9.64E-02 | -0.170 | 2.61E-08 | **1.58E-07** | 0.007 | 7.87E-01 | 8.31E-01 |
| Q99497 | PARK7 | 0.289 | 4.14E-03 | 1.06E-01 | -0.247 | 5.14E-04 | **1.04E-03** | 0.436 | 3.57E-09 | **8.79E-08** |
| P02452 | COL1A1 | -0.164 | 4.30E-03 | 1.06E-01 | -0.248 | 2.23E-08 | **1.40E-07** | 0.062 | 1.17E-01 | 1.95E-01 |
| O75594 | PGLYRP1 | 0.266 | 5.91E-03 | 1.25E-01 | 0.008 | 9.19E-01 | 9.34E-01 | 0.189 | 1.25E-02 | **3.64E-02** |
| O95185 | UNC5C | -0.202 | 5.38E-03 | 1.25E-01 | -0.391 | 1.94E-11 | **4.77E-10** | 0.084 | 8.41E-02 | 1.51E-01 |
| Q9HCK4 | ROBO2 | -0.270 | 5.85E-03 | 1.25E-01 | -0.504 | 8.64E-11 | **1.44E-09** | 0.107 | 1.06E-01 | 1.79E-01 |
| P80370 | DLK-1 | -0.216 | 5.96E-03 | 1.25E-01 | -0.335 | 8.04E-08 | **4.24E-07** | 0.078 | 1.77E-01 | 2.69E-01 |
| O60259 | hK8 | -0.196 | 6.03E-03 | 1.25E-01 | -0.190 | 2.35E-03 | **3.99E-03** | -0.045 | 3.94E-01 | 4.93E-01 |
| O43155 | FLRT2 | -0.183 | 6.23E-03 | 1.25E-01 | -0.328 | 3.82E-09 | **3.68E-08** | 0.080 | 9.13E-02 | 1.60E-01 |
| Q9BYH1 | SEZ6L | -0.058 | 7.01E-03 | 1.37E-01 | -0.095 | 1.40E-09 | **1.69E-08** | 0.015 | 2.46E-01 | 3.42E-01 |
| Q6NW40 | RGMB | -0.165 | 7.43E-03 | 1.37E-01 | -0.290 | 3.37E-10 | **5.09E-09** | 0.054 | 1.68E-01 | 2.58E-01 |
| P05067 | APP | -0.089 | 7.34E-03 | 1.37E-01 | -0.139 | 7.50E-09 | **5.94E-08** | 0.019 | 3.63E-01 | 4.66E-01 |
| Q96PQ0 | SORCS2 | -0.075 | 8.04E-03 | 1.41E-01 | -0.119 | 1.79E-09 | **2.09E-08** | 0.043 | 1.50E-02 | **4.27E-02** |
| Q9BZR6 | RTN4R | -0.146 | 8.09E-03 | 1.41E-01 | -0.191 | 1.98E-05 | **5.47E-05** | 0.017 | 6.45E-01 | 7.15E-01 |
| Q9NQ88 | TIGAR | 0.095 | 9.51E-03 | 1.58E-01 | -0.114 | 4.00E-04 | **8.48E-04** | 0.164 | 1.94E-08 | **4.03E-07** |
| P50591 | TRAIL | -0.131 | 9.81E-03 | 1.59E-01 | -0.217 | 1.04E-06 | **4.10E-06** | 0.034 | 3.78E-01 | 4.80E-01 |
| Q9UHF1 | EGFL7 | -0.184 | 1.02E-02 | 1.62E-01 | -0.352 | 4.37E-08 | **2.44E-07** | 0.038 | 4.76E-01 | 5.75E-01 |
| Q6UX15 | LAYN | -0.223 | 1.07E-02 | 1.65E-01 | -0.389 | 4.85E-09 | **4.30E-08** | 0.107 | 5.93E-02 | 1.17E-01 |
| O95727 | CRTAM | -0.132 | 1.11E-02 | 1.67E-01 | -0.220 | 2.15E-06 | **7.63E-06** | 0.017 | 6.51E-01 | 7.19E-01 |
| P16234 | DGF-R-alph | -0.168 | 1.14E-02 | 1.69E-01 | -0.279 | 3.92E-07 | **1.76E-06** | 0.030 | 5.09E-01 | 6.02E-01 |
| O95750 | FGF-19 | -0.220 | 1.17E-02 | 1.69E-01 | -0.379 | 5.27E-08 | **2.87E-07** | 0.000 | 9.94E-01 | 9.96E-01 |
| P78423 | CX3CL1 | -0.177 | 1.25E-02 | 1.69E-01 | -0.446 | 1.23E-14 | **8.19E-13** | 0.166 | 9.95E-04 | **5.21E-03** |
| Q8TAD2 | IL-17D | 0.170 | 1.23E-02 | 1.69E-01 | -0.049 | 3.91E-01 | 4.32E-01 | 0.143 | 7.51E-03 | **2.56E-02** |
| Q8NEV9,Q | IL-27 | 0.173 | 1.32E-02 | 1.69E-01 | -0.021 | 7.12E-01 | 7.47E-01 | 0.131 | 9.34E-03 | **2.97E-02** |
| P80162 | CXCL6 | 0.237 | 1.24E-02 | 1.69E-01 | -0.010 | 8.97E-01 | 9.17E-01 | 0.153 | 3.41E-02 | 7.62E-02 |
| Q14508 | WFDC2 | -0.086 | 1.28E-02 | 1.69E-01 | -0.168 | 4.54E-09 | **4.08E-08** | 0.048 | 4.10E-02 | 8.71E-02 |
| Q96B86 | RGMA | -0.091 | 1.31E-02 | 1.69E-01 | -0.181 | 7.58E-11 | **1.33E-09** | 0.046 | 4.64E-02 | 9.61E-02 |
| P10147 | CCL3 | 0.173 | 1.41E-02 | 1.77E-01 | -0.161 | 4.07E-03 | **6.65E-03** | 0.331 | 1.61E-10 | **5.11E-09** |
| P18065 | IGFBP-2 | -0.140 | 1.45E-02 | 1.79E-01 | -0.229 | 1.10E-07 | **5.52E-07** | 0.084 | 2.48E-02 | 6.20E-02 |
| O43505 | B4GAT1 | -0.082 | 1.49E-02 | 1.80E-01 | -0.125 | 9.39E-08 | **4.81E-07** | 0.031 | 1.31E-01 | 2.13E-01 |
| Q9BUD6 | SPON2 | -0.115 | 1.52E-02 | 1.81E-01 | -0.126 | 1.37E-03 | **2.41E-03** | -0.065 | 7.30E-02 | 1.37E-01 |
| P50579 | MetAP 2 | 0.117 | 1.69E-02 | 1.91E-01 | -0.147 | 2.99E-05 | **7.82E-05** | 0.203 | 3.01E-08 | **5.97E-07** |
| O75509 | TNFRSF21 | -0.065 | 1.67E-02 | 1.91E-01 | -0.107 | 8.14E-09 | **6.29E-08** | 0.022 | 1.80E-01 | 2.72E-01 |
| P41271 | NBL1 | 0.091 | 1.67E-02 | 1.91E-01 | 0.207 | 1.04E-08 | **7.59E-08** | -0.035 | 2.50E-01 | 3.46E-01 |
| P31431 | SDC4 | 0.138 | 1.95E-02 | 2.14E-01 | -0.319 | 5.37E-11 | **1.11E-09** | 0.447 | 1.31E-21 | **2.90E-19** |
| Q8NFT8 | DNER | -0.049 | 2.06E-02 | 2.14E-01 | -0.090 | 1.36E-08 | **9.23E-08** | 0.041 | 9.64E-04 | **5.12E-03** |
| Q9Y275 | TNFSF13B | -0.195 | 2.01E-02 | 2.14E-01 | -0.391 | 1.07E-08 | **7.65E-08** | 0.199 | 1.59E-03 | **7.61E-03** |
| P21246 | PTN | -0.091 | 2.05E-02 | 2.14E-01 | 0.011 | 7.60E-01 | 7.92E-01 | -0.073 | 2.99E-02 | 7.08E-02 |
| Q9UNE0 | EDAR | 0.089 | 1.97E-02 | 2.14E-01 | -0.001 | 9.87E-01 | 9.87E-01 | 0.049 | 9.98E-02 | 1.72E-01 |
| Q2TAL6 | VWC2 | -0.183 | 2.10E-02 | 2.15E-01 | -0.439 | 8.65E-12 | **2.40E-10** | 0.114 | 3.32E-02 | 7.56E-02 |
| Q16627 | CCL14 | 0.196 | 2.20E-02 | 2.22E-01 | 0.002 | 9.82E-01 | 9.83E-01 | 0.136 | 3.34E-02 | 7.57E-02 |
| Q96PL1 | SCGB3A2 | 0.136 | 2.30E-02 | 2.28E-01 | -0.038 | 5.05E-01 | 5.47E-01 | 0.129 | 1.05E-02 | **3.27E-02** |
| P36222 | CHI3L1 | -0.056 | 2.40E-02 | 2.33E-01 | 0.023 | 2.25E-01 | 2.60E-01 | -0.012 | 5.24E-01 | 6.15E-01 |
| Q8TEU8 | WFIKKN2 | -0.226 | 2.41E-02 | 2.33E-01 | -0.290 | 2.01E-04 | **4.56E-04** | 0.045 | 5.27E-01 | 6.15E-01 |
| P25116 | PAR-1 | 0.394 | 2.63E-02 | 2.50E-01 | -0.017 | 9.13E-01 | 9.30E-01 | 0.360 | 1.08E-02 | **3.35E-02** |
| O94779 | CNTN5 | -0.162 | 2.89E-02 | 2.60E-01 | -0.404 | 3.23E-12 | **1.07E-10** | 0.144 | 2.63E-03 | **1.09E-02** |
| Q969Z4 | RELT | -0.195 | 2.78E-02 | 2.60E-01 | -0.472 | 6.50E-12 | **1.97E-10** | 0.170 | 5.45E-03 | **2.00E-02** |
| P24387 | CRHBP | -0.136 | 2.82E-02 | 2.60E-01 | -0.121 | 9.56E-03 | **1.48E-02** | -0.049 | 2.85E-01 | 3.87E-01 |
| Q9UBT3 | Dkk-4 | -0.152 | 2.87E-02 | 2.60E-01 | -0.272 | 1.68E-06 | **6.12E-06** | 0.019 | 7.04E-01 | 7.56E-01 |
| Q9NP99 | TREM1 | 0.115 | 3.08E-02 | 2.70E-01 | -0.282 | 1.09E-06 | **4.21E-06** | 0.363 | 1.07E-13 | **5.09E-12** |
| Q92520 | FAM3C | -0.028 | 3.06E-02 | 2.70E-01 | -0.016 | 1.67E-01 | 1.97E-01 | 0.006 | 5.53E-01 | 6.36E-01 |
| P13688 | CEACAM1 | 0.146 | 3.23E-02 | 2.79E-01 | -0.025 | 6.70E-01 | 7.07E-01 | 0.055 | 2.95E-01 | 3.97E-01 |
| Q8TCZ2 | CD99L2 | -0.081 | 3.42E-02 | 2.88E-01 | -0.156 | 7.42E-09 | **5.94E-08** | 0.050 | 4.32E-02 | 9.12E-02 |
| P15151 | PVR | -0.170 | 3.41E-02 | 2.88E-01 | -0.399 | 2.72E-09 | **2.89E-08** | 0.109 | 5.91E-02 | 1.17E-01 |
| O00548 | DLL1 | -0.161 | 3.54E-02 | 2.94E-01 | -0.375 | 7.37E-09 | **5.94E-08** | 0.089 | 9.82E-02 | 1.70E-01 |
| P12544 | GZMA | -0.162 | 3.65E-02 | 2.99E-01 | -0.231 | 4.15E-04 | **8.71E-04** | 0.036 | 5.42E-01 | 6.30E-01 |
| Q7Z5A7 | FAM19A5 | -0.058 | 3.96E-02 | 3.18E-01 | -0.101 | 1.17E-08 | **8.22E-08** | 0.023 | 2.12E-01 | 3.08E-01 |
| O43240 | KLK10 | -0.198 | 3.95E-02 | 3.18E-01 | -0.264 | 8.99E-04 | **1.69E-03** | 0.040 | 5.67E-01 | 6.47E-01 |
| Q8NBP7 | PCSK9 | -0.143 | 4.08E-02 | 3.23E-01 | -0.257 | 4.85E-06 | **1.56E-05** | 0.031 | 5.15E-01 | 6.07E-01 |
| Q9H5V8 | CDCP1 | 0.158 | 4.36E-02 | 3.38E-01 | -0.056 | 3.64E-01 | 4.05E-01 | 0.126 | 1.90E-02 | 5.16E-02 |
| P35318 | ADM | -0.120 | 4.38E-02 | 3.38E-01 | -0.270 | 9.93E-09 | **7.42E-08** | 0.081 | 4.81E-02 | 9.86E-02 |
| Q9NZ53 | PODXL2 | -0.074 | 4.49E-02 | 3.43E-01 | -0.124 | 1.66E-06 | **6.10E-06** | 0.022 | 3.60E-01 | 4.64E-01 |
| O60462 | NRP2 | -0.119 | 4.67E-02 | 3.53E-01 | -0.253 | 5.38E-07 | **2.29E-06** | 0.089 | 3.56E-02 | 7.80E-02 |
| P57087 | JAM-B | -0.095 | 4.81E-02 | 3.59E-01 | -0.213 | 4.38E-09 | **3.99E-08** | 0.058 | 7.42E-02 | 1.38E-01 |
| O14793 | GDF-8 | -0.123 | 4.98E-02 | 3.65E-01 | -0.245 | 3.35E-06 | **1.14E-05** | 0.040 | 3.73E-01 | 4.75E-01 |

Table shows the effect (beta coefficient), *p* values and the resulting q values after applying FDR post-hoc correction of each of the CSF protein abundance that differed between DLB and CON (blue) or AD (purple) or AD and CON (red). Significant proteins (q <0.05) are highlighted in bold.
