## Supplementary figures for "CSF proteome profiling reveals highly specific biomarkers for dementia with Lewy bodies"

a

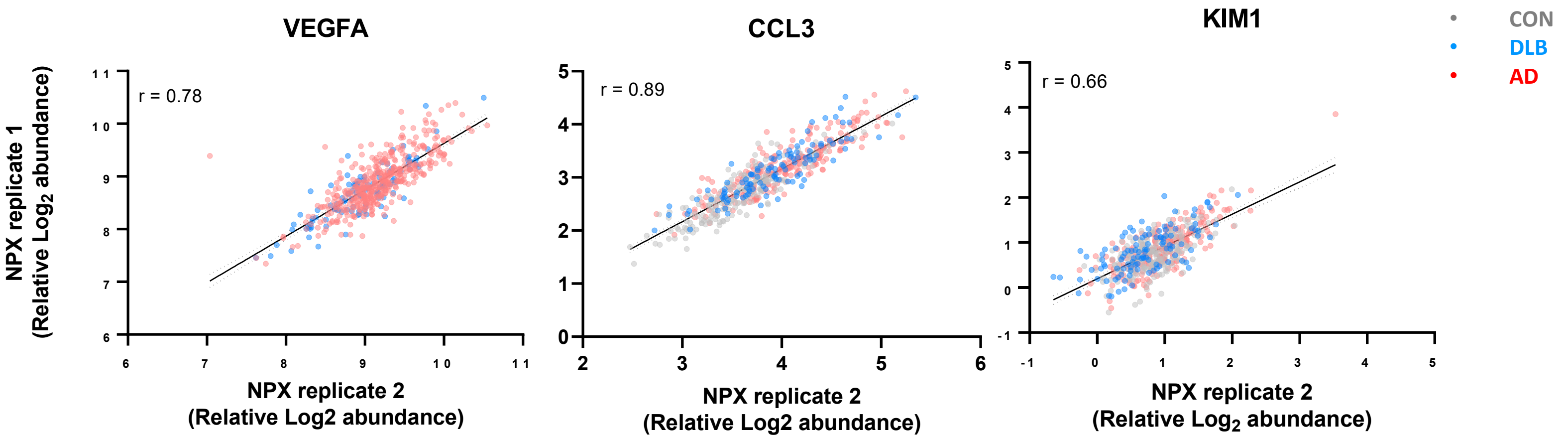


**Supplementary figure 1. Correlations between protein replicates measured through different PEA panels. a**, Scatter plots show the correlation of the NPX values (protein abundance) between the protein replicates that showed significant differences across the groups of interest in the complete cohort (n=534). Regression line and 95% confident intervals are depicted. Insert indicate the spearman correlation coefficient. DLB, Dementia with Lewy Bodies; CON, cognitively unimpaired controls; AD, Alzheimer's disease

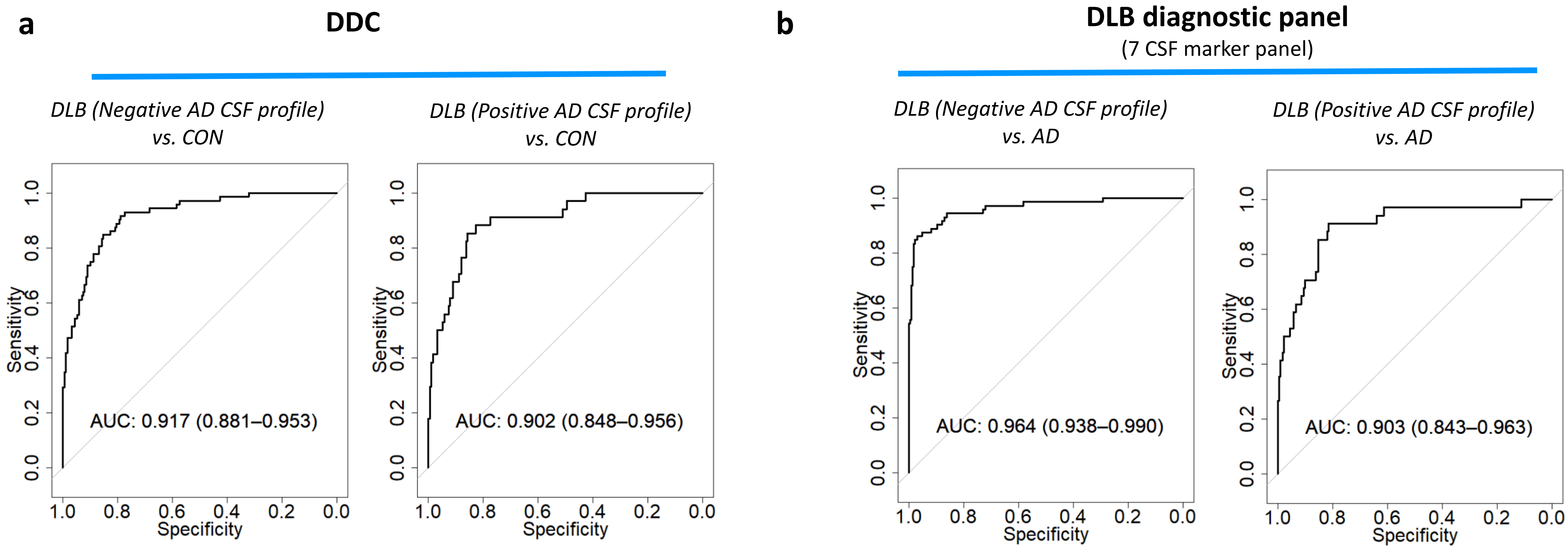


**Supplementary figure 2. Additional sensitivity analysis for the CSF biomarker panels discriminating DLB from controls or AD a)** Receiver operating characteristic (ROC) curves showed that DDC (a) or the CSF DLB-diagnostic panel (b) could discriminate DLB from controls or AD with similar performance independent of the presence of AD co-pathology in DLB patients (as measured by positive or negative AD CSF biomarker profiles). Inserts outline corresponding AUC and 95% CI. DLB, dementia with Lewy bodies; CON, cognitively unimpaired controls; AD, Alzheimer's disease;

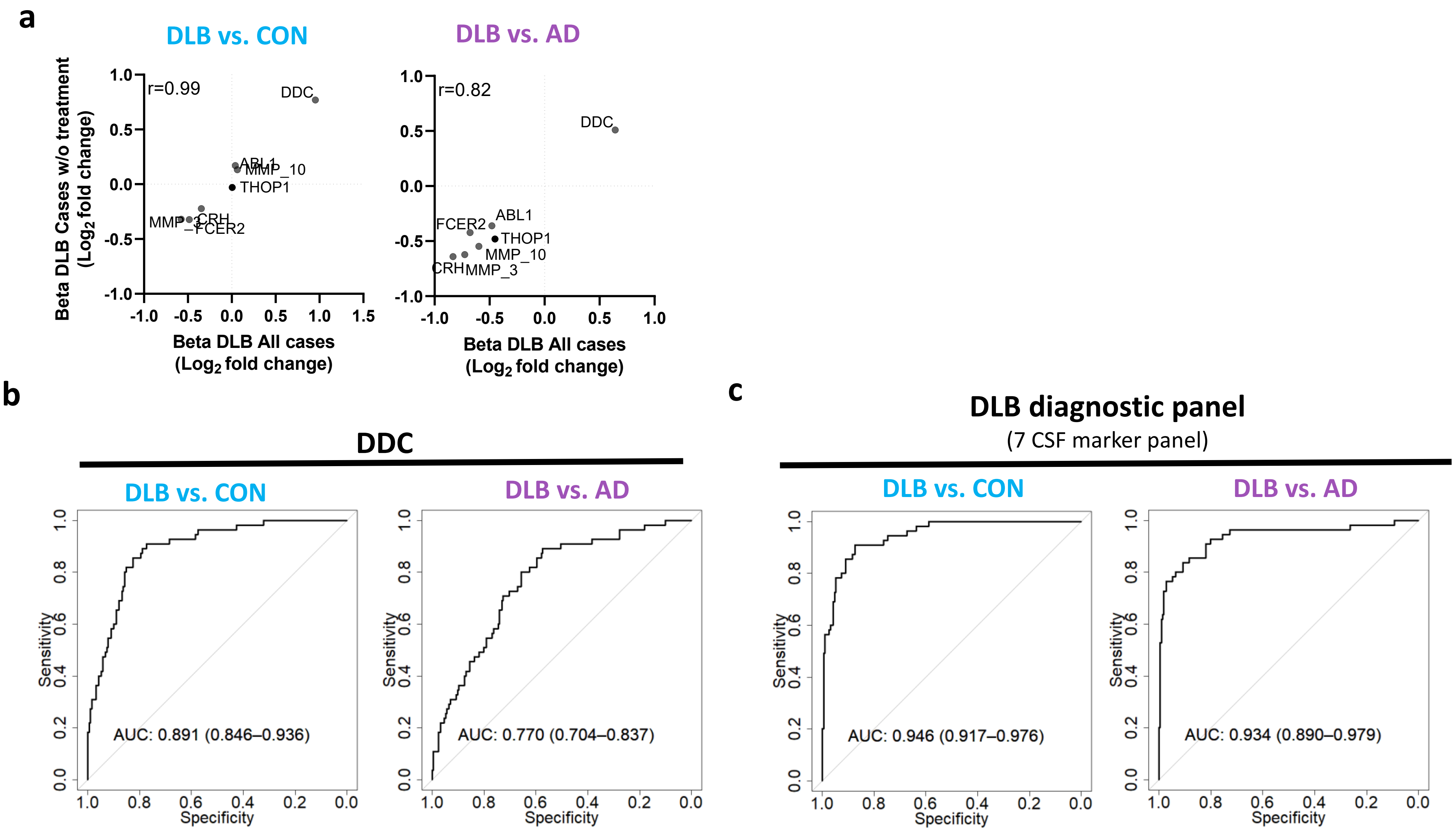


**Supplementary figure 3. Additional sensitivity analysis for the CSF biomarker panel discriminating DLB from controls or AD in the discovery cohorts including only DLB patients that are not undergoing parkinsonian related treatment**. **a**) Scatter plots show the correlation between the beta-coefficients obtained in the complete discovery cohorts to those obtained when only including patients that did not have parkinsonian related medication (DLB=55, CON=190, AD=216). Insert indicate the spearman correlation coefficient. **b-c)**Receiver operating characteristic (ROC) curves depicting the performance of (b) DDC or (c) the CSF DLB-diagnostic panel in the comparison between DLB (n=55) and controls (n=190) or AD (n=216) in the discovery cohorts after including patients that are not undergoing any parkinsonian related treatment. Inserts outline corresponding AUC and 95% CI. DLB, dementia with Lewy bodies; CON, cognitively unimpaired controls; AD, Alzheimer's disease.

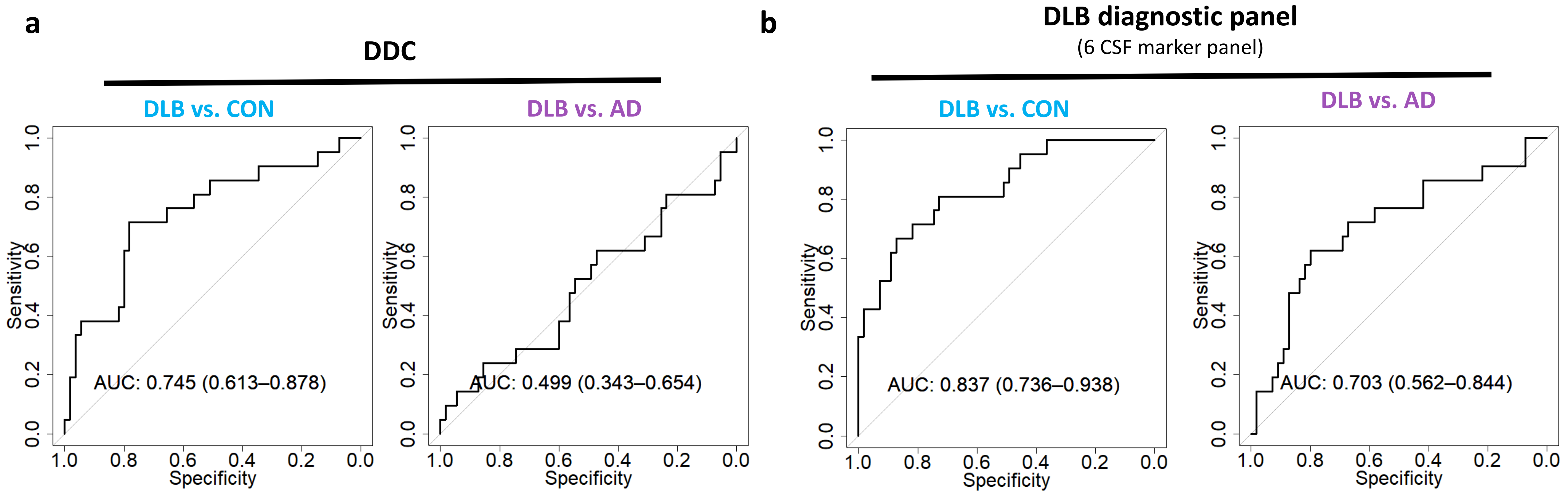


**Supplementary figure 4. Additional sensitivity analysis for the CSF biomarker panel discriminating DLB from controls or AD in the second validation cohort including only DLB patients with an abnormal DAT scan.** Receiver operating characteristic (ROC) curves depicting the performance of (a) DDC or (b) the CSF DLB-diagnostic panel in the comparison between DLB (n=21) and controls (n=55) or AD (n=55) in the second validation cohort after including only DLB cases with abnormal DaTScan. Inserts outline corresponding AUC and 95% CI. DLB, dementia with Lewy bodies; CON, cognitively unimpaired controls; AD, Alzheimer's disease.
