## Supplementary tables for "CSF proteome profiling reveals highly specific biomarkers for dementia with Lewy bodies"

**Supplementary Table 1. Biomarkers supporting DLB diagnosis across cohorts**

| | Discovery cohort | Validation cohort 1 | Validation cohort 2 | Autopsy cohort |
|---|---|---|---|---|
| **Autopsy confirmed, n (%)** | 14 (13%) | 0 (0%) | 0 (0%) | 17 (100%) |
| **Clinical based diagnosis, n (%)** | 95 (87%) | 54 (100%) | 55 (100%) | na |
| **DAT-SPECT, n abnormal (%)** | 23 (24%) | 19 (35%) | 21 (38%) | na |
| **EEG, n abnormal (%)** | 41 (43%) | 28 (51%) | na | na |
| **No autopsy or supporting biomarkers available, n (%)** | 48 (44%) | 18 (33%) | 27 (49%) | na |

Abbreviations: DAT-SPECT: FPCIT single-photon emission computed tomography; EEG: electroencephalography.

**Supplementary Table 2. Proteins that passed the quality control and were included in the analysis**

| Name | Abbreviaton | Uniprot | Olink panel(s) | CSF Detectavility |
|---|---|---|---|---|
| C-C motif chemokine 24 | CCL24 | O00175 | CVD III | 91.22% |
| Galectin-9 | Gal-9 | O00182 | CVD II | 100.00% |
| Galectin-8 | gal-8 | O00214 | Neuro | 99.90% |
| Tumor necrosis factor receptor superfamily member 10A | TNFRSF10A | O00220 | CVD II, Cell-Reg | 100%; 100% |
| Signal-regulatory protein beta-1 | SIRPB1 | O00241 | Dev | 100.00% |
| Agouti-related protein | AGRP | O00253 | CVD II | 100.00% |
| DNA fragmentation factor subunit alpha | DFFA | O00273 | IR | 100.00% |
| Osteoprotegerin | OPG | O00300 | Inf, CVD III | 100%; 100% |
| Matrilin-2 | MATN2 | O00339 | Dev | 100.00% |
| GDNF family receptor alpha-2 | GFRA2 | O00451 | Cell-Reg | 100.00% |
| Neural cell adhesion molecule L1-like protein | CHL1 | O00533 | C-Met | 100.00% |
| Delta-like protein 1 | DLL1 | O00548 | Onc II | 100.00% |
| C-C motif chemokine 21 | CCL21 | O00585 | Dev | 96.79% |
| Podocalyxin | PODXL | O00592 | Onc II | 99.69% |
| Protein CYR61 | CYR61 | O00622 | Onc II | 100.00% |
| Neurocan core protein | NCAN | O14594 | Neuro | 100.00% |
| C-X-C motif chemokine 11 | CXCL11 | O14625 | Inf | 99.90% |
| TNF-related apoptosis-inducing ligand receptor 2 | TRAIL-R2 | O14763 | CVD II | 100.00% |
| Tripeptidyl-peptidase 1 | TPP1 | O14773 | Dev | 100.00% |
| Growth/differentiation factor 8 | GDF-8 | O14793 | Neuro | 100.00% |
| Tumor necrosis factor receptor superfamily member 10C | TNFRSF10C | O14798 | CVD III | 100.00% |
| Secretory carrier-associated membrane protein 3 | SCAMP3 | O14828 | Onc II | 100.00% |
| Tumor necrosis factor receptor superfamily member 13B | TNFRSF13B | O14836 | CVD II | 100.00% |
| Protein Wnt-9a | WNT9A | O14904 | Cell-Reg | 90.49% |
| Protocadherin-17 | PCDH17 | O14917 | Cell-Reg | 100.00% |
| Plexin-B2 | PLXNB2 | O15031 | C-Met | 100.00% |
| Angiopoietin-2 | ANGPT2 | O15123 | Met | 100.00% |
| Ephrin type-B receptor 6 | EPHB6 | O15197 | Neuro | 99.90% |
| Matrilin-3 | MATN3 | O15232 | Neuro | 100.00% |
| C-C motif chemokine 25 | CCL25 | O15444 | Inf | 97.52% |
| Toll-like receptor 3 | TLR3 | O15455 | Onc II | 100.00% |
| C-C motif chemokine 16 | CCL16 | O15467 | CVD III | 100.00% |
| Leucine-rich repeat transmembrane protein FLRT2 | FLRT2 | O43155 | Neuro | 99.90% |
| Plexin-B1 | PLXNB1 | O43157 | Neuro | 100.00% |
| Kallikrein-10 | KLK10 | O43240 | Met | 100.00% |
| Kunitz-type protease inhibitor 1 | SPINT1 | O43278 | Dev | 100.00% |
| Kunitz-type protease inhibitor 2 | SPINT2 | O43291 | Dev | 100.00% |
| Cochlin | COCH | O43405 | Dev | 100.00% |
| Beta-1,4-glucuronyltransferase 1 | B4GAT1 | O43505 | Dev | 100.00% |
| Tumor necrosis factor (Ligand) superfamily, member 12 | TWEAK | O43508 | Inf | 100.00% |
| Bcl-2-like protein 11 isoform BimL | BCL2L11 | O43521-2 | Cell-Reg | 100.00% |
| Tumor necrosis factor ligand superfamily member 14 | TNFSF14 | O43557 | Inf | 96.59% |
| Carbonic anhydrase 12 | CA12 | O43570 | OD | 99.48% |
| Protein sprouty homolog 2 | SPRY2 | O43597 | IR | 100.00% |
| Integral membrane protein 2A | ITM2A | O43736 | IR | 100.00% |
| Angiopoietin-related protein 7 | ANGPTL7 | O43827 | Met | 100.00% |
| EGF-like repeat and discoidin I-like domain-containing protein 3 | EDIL3 | O43854 | OD | 99.90% |
| Xaa-Pro aminopeptidase 2 | XPNPEP2 | O43895 | Onc II | 97.52% |
| Vascular endothelial growth factor D | VEGFD | O43915 | CVD II, Cell-Reg | 100%; 99,79% |
| C-X-C motif chemokine 13 | CXCL13 | O43927 | Onc II | 100.00% |
| Heparan-sulfate 6-O-sulfotransferase 1 | HS6ST1 | O60243 | Cell-Reg | 100.00% |
| Kallikrein-8 | hK8 | O60259 | Onc II | 100.00% |
| Neuropilin-2 | NRP2 | O60462 | Neuro | 99.69% |
| GDNF family receptor alpha-3 | GDNFR-alpha-3 | O60609 | Neuro | 100.00% |
| Cathepsin L2 | CTSV | O60911 | Onc II | 100.00% |
| Low affinity immunoglobulin gamma Fc region receptor III-B | FCGR3B | O75015 | C-Met | 100.00% |
| Leukocyte immunoglobulin-like receptor subfamily B member 5 | LILRB5 | O75023 | C-Met | 99.28% |
| Immunoglobulin superfamily member 3 | IGSF3 | O75054 | Cell-Reg | 100.00% |
| Disintegrin and metalloproteinase domain-containing protein 23 | ADAM 23 | O75077 | Neuro | 100.00% |
| ICOS ligand | ICOSLG | O75144 | Onc II | 100.00% |
| Semaphorin-7A | SEMA7A | O75326 | Dev | 100.00% |
| Ectonucleoside triphosphate diphosphohydrolase 6 | ENTPD6 | O75354 | OD, Cell-Reg | 100%; 99,79% |
| Ectonucleoside triphosphate diphosphohydrolase 5 | ENTPD5 | O75356 | Met | 98.97% |
| Tumor necrosis factor receptor superfamily member 21 | TNFRSF21 | O75509 | Neuro | 100.00% |
| Peptidoglycan recognition protein 1 | PGLYRP1 | O75594 | CVD III | 100.00% |
| Tumor necrosis factor ligand superfamily member 13 | TNFSF13 | O75888 | Onc II | 100.00% |
| N(G),N(G)-dimethylarginine dimethylaminohydrolase 1 (DDAH1) | DDAH1 | O94760 | Cell-Reg | 100.00% |
| Contactin-5 | CNTN5 | O94779 | Neuro | 100.00% |
| Dickkopf-related protein 1 | Dkk-1 | O94907 | CVD II | 100.00% |
| Vesicle-associated membrane protein 5 | VAMP5 | O95183 | Cell-Reg | 89.45% |
| Netrin receptor UNC5C | UNC5C | O95185 | Neuro | 99.90% |
| WNT1-inducible-signaling pathway protein 1 | WISP-1 | O95388 | Onc II | 100.00% |
| Tumor necrosis factor receptor superfamily member 6B | TNFRSF6B | O95407 | Onc II | 100.00% |
| Apolipoprotein M | APOM | O95445 | C-Met | 98.76% |
| Neuronal pentraxin receptor | NPTXR | O95502 | Met | 100.00% |
| NAD kinase | DK | O95544 | Met | 99.90% |
| Follistatin-related protein 3 | FSTL3 | O95633 | Dev | 100.00% |
| Synaptosomal-associated protein 29 | SP29 | O95721 | Dev | 100.00% |
| Cytotoxic and regulatory T-cell molecule | CRTAM | O95727 | Neuro | 92.98% |
| Fibroblast growth factor 19 | FGF-19 | O95750 | Inf | 100.00% |
| Angiopoietin-related protein 1 | ANGPTL1 | O95841 | Met | 100.00% |
| CD160 antigen | CD160 | O95971 | Onc II | 99.90% |
| Interleukin-18-binding protein | IL-18BP | O95998 | CVD III | 100.00% |
| Superoxide dismutase (Cu-Zn) | SOD1 | P00441 | C-Met | 100.00% |
| Tyrosine-protein kinase ABL1 | ABL1 | P00519 | Onc II | 100.00% |
| Epidermal growth factor receptor | EGFR | P00533 | CVD III | 100.00% |
| Urokinase-type plasminogen activator | uPA | P00749 | Inf, CVD III | 100%; 100% |
| Tissue-type plasminogen activator | t-PA | P00750 | CVD III | 100.00% |
| Adenosine Deaminase | ADA | P00813 | Inf | 100.00% |
| Carbonic anhydrase 2 | CA2 | P00918 | Dev | 91.11% |
| Serine protease inhibitor Kazal-type 1 | SPINK1 | P00995 | Dev | 100.00% |
| Metalloproteinase inhibitor 1 | TIMP1 | P01033 | C-Met | 100.00% |
| Cystatin-C | CST3 | P01034 | C-Met | 100.00% |
| Platelet-derived growth factor subunit B | PDGF subunit B | P01127 | CVD II | 100.00% |
| Low-density lipoprotein receptor | LDL receptor | P01130 | CVD III | 100.00% |
| Transforming growth factor alpha | TGF-alpha | P01135 | Inf, Onc II | 100%; 100% |
| Latency-associated peptide transforming growth factor beta-1 | LAP TGF-beta-1 | P01137 | Inf | 100.00% |
| Beta-nerve growth factor | Beta-NGF | P01138 | Inf, Neuro | 100%; 99,9% |
| Glycoprotein hormones alpha chain | CGA | P01215 | Dev | 100.00% |
| Growth hormone | GH | P01241 | CVD II | 98.76% |
| Calcitonin | CALCA | P01258 | OD | 99.90% |
| Pancreatic prohormone | PPY | P01298 | Onc II | 100.00% |
| TNF-beta | TNFB | P01374 | Inf | 98.24% |
| Interleukin-2 receptor subunit alpha | IL2-RA | P01589 | CVD III | 99.48% |
| T-cell surface glycoprotein CD4 | CD4 | P01730 | CVD II | 100.00% |
| Myoglobin | MB | P02144 | CVD III | 100.00% |
| Collagen alpha-1(I) chain | COL1A1 | P02452 | CVD III | 100.00% |
| Collagen alpha-1(IV) chain | COL4A1 | P02462 | Cell-Reg | 100.00% |
| Protein AMBP | AMBP | P02760 | CVD II | 100.00% |
| C-X-C motif chemokine 10 | CXCL10 | P02778 | Inf | 100.00% |
| Transferrin receptor protein 1 | TR | P02786 | CVD III | 99.79% |
| Angiogenin | ANG | P03950 | C-Met | 100.00% |
| Coagulation factor XI | F11 | P03951 | C-Met | 100.00% |
| Matrix metalloproteinase-1 | MMP-1 | P03956 | Inf | 100.00% |
| Tissue alpha-L-fucosidase | FUCA1 | P04066 | Dev | 100.00% |
| Vitamin K-dependent protein C | PROC | P04070 | C-Met | 100.00% |
| Cystatin-B | CSTB | P04080 | CVD III | 100.00% |
| Platelet-derived growth factor subunit A | PDGF subunit A | P04085 | CVD III | 100.00% |
| Superoxide dismutase (Mn), mitochondrial | SOD2 | P04179 | CVD II | 100.00% |
| Thy-1 membrane glycoprotein | THY 1 | P04216 | Neuro | 100.00% |
| HLA class II histocompatibility antigen gamma chain | CD74 | P04233 | Dev | 99.38% |
| von Willebrand factor | vWF | P04275 | CVD III | 97.52% |
| Receptor tyrosine-protein kinase erbB-2 | ErbB2/HER2 | P04626 | Onc II | 100.00% |
| Heat shock 27 kDa protein | HSP 27 | P04792 | CVD II | 100.00% |
| Amyloid beta A4 protein | APP | P05067 | Dev | 100.00% |
| Integrin beta-2 | ITGB2 | P05107 | CVD III | 100.00% |
| Plasminogen activator inhibitor 1 | PAI | P05121 | CVD III | 100.00% |
| Plasma serine protease inhibitor | SERPINA5 | P05154 | C-Met | 100.00% |
| Interleukin-6 | IL6 | P05231 | Inf, Onc II, CVD II, IR | 100%; 100%; 100%; 100% |
| Intercellular adhesion molecule 1 | ICAM1 | P05362 | C-Met | 100.00% |
| Lithostathine-1-alpha | REG1A | P05451 | C-Met | 100.00% |
| Thyroxine-binding globulin | SERPINA7 | P05543 | C-Met | 99.59% |
| Integrin beta-1 | ITGB1 | P05556 | Dev | 100.00% |
| T-cell surface glycoprotein CD5 | CD5 | P06127 | Inf | 100.00% |
| Complement C2 | C2 | P06681 | C-Met | 100.00% |

| | | | | |
|---|---|---|---|---|
| Low affinity immunoglobulin epsilon Fc receptor | FCER2 | P06734 | Dev | 100.00% |
| Integrin alpha-V | ITGAV | P06756 | Onc II | 100.00% |
| Corticoliberin | CRH | P06850 | OD | 99.59% |
| Lipoprotein lipase | LPL | P06858 | CVD II | 99.90% |
| Thrombomodulin TM | | P07204 | CVD II | 100.00% |
| Protein disulfide-isomerase | P4HB | P07237 | Dev | 95.14% |
| Cathepsin D | CTSD | P07339 | CVD III | 100.00% |
| Platelet glycoprotein Ib alpha chain | GP1BA | P07359 | C-Met | 90.70% |
| Carbonic anhydrase 3 | CA3 | P07451 | C-Met | 92.77% |
| Trypsin-2 | PRSS2 | P07478 | C-Met | 99.59% |
| Decorin | DCN | P07585 | CVD II | 100.00% |
| Cathepsin L1 | CTSL1 | P07711 | CVD II | 100.00% |
| Tyrosine-protein kinase Yes | YES1 | P07947 | OD | 99.79% |
| Tyrosine-protein kinase Lyn | LYN | P07948 | Onc II | 98.24% |
| Proto-oncogene tyrosine-protein kinase receptor Ret | RET | P07949 | Onc II | 100.00% |
| Insulin-like growth factor 1 receptor | IGF1R | P08069 | Onc II | 100.00% |
| Beta-glucuronidase | GUSB | P08236 | Dev | 100.00% |
| Matrix metalloproteinase-2 | MMP-2 | P08253 | CVD III | 100.00% |
| Matrix metalloproteinase-3 | MMP-3 | P08254 | CVD III | 100.00% |
| Neprilysin | NEP | P08473 | Neuro | 99.69% |
| Hepatocyte growth factor receptor | MET | P08581 | C-Met | 100.00% |
| Vimentin | VIM | P08670 | Onc II | 94.63% |
| Coagulation factor VII | F7 | P08709 | C-Met | 88.43% |
| Insulin-like growth factor-binding protein 1 | IGFBP-1 | P08833 | CVD III | 100.00% |
| Interleukin-6 receptor subunit alpha | IL-6RA | P08887 | CVD III | 100.00% |
| Gamma-enolase | ENO2 | P09104 | Met | 100.00% |
| Matrix metalloproteinase-7 | MMP-7 | P09237 | CVD II | 100.00% |
| Matrix metalloproteinase-10 | MMP-10 | P09238 | Inf | 99.79% |
| CD48 antigen | CD48 | P09326 | Onc II | 100.00% |
| C-X-C motif chemokine 1 | CXCL1 | P09341 | Inf,CVD II | 100%; 100% |
| Galectin-1 | Gal-1 | P09382 | Onc II | 100.00% |
| Dihydropteridine reductase | QDPR | P09417 | Met | 100.00% |
| SPARC | SPARC | P09486 | Onc II | 100.00% |
| Heme oxygenase 1 | HO-1 | P09601 | CVD II | 99.90% |
| Macrophage colony-stimulating factor 1 | CSF-1 | P09603 | Inf | 100.00% |
| Platelet-derived growth factor receptor beta | PDGFRB | P09619 | Dev | 100.00% |
| Pro-cathepsin H | CTSH | P09668 | Met | 100.00% |
| Tumor-associated calcium signal transducer 2 | TACSTD2 | P09758 | Cell-Reg | 100.00% |
| Furin | FUR | P09958 | Onc II | 100.00% |
| Cryptic protein | CFC1 | P0CG37 | Cell-Reg | 100.00% |
| Ig lambda-2 chain C regions | IGLC2 | P0DOY2 | C-Met | 100.00% |
| Granzyme B | GZMB | P10144 | Onc II | 85.74% |
| Interleukin-8 | IL-8 | P10145 | Inf | 100.00% |
| C-C motif chemokine 3 | CCL3 | P10147 | Inf, CVD II | 100%; 100% |
| Osteopontin | OPN | P10451 | CVD III | 100.00% |
| Receptor-type tyrosine-protein phosphatase F | PTPRF | P10586 | Dev | 100.00% |
| Tissue factor pathway inhibitor | TFPI | P10646 | CVD III | 100.00% |
| Mast/stem cell growth factor receptor Kit | KIT | P10721 | C-Met | 100.00% |
| Mannose-binding protein C | MBL2 | P11226 | C-Met | 99.48% |
| Cation-independent mannose-6-phosphate receptor | IGF2R | P11717 | Dev | 100.00% |
| Fatty acid-binding protein, intestinal (FABP2) | FABP2 | P12104 | CVD II | 97.00% |
| Low affinity immunoglobulin gamma Fc region receptor II-a | FCGR2A | P12318 | C-Met | 99.38% |
| Granzyme A | GZMA | P12544 | Neuro | 99.90% |
| Bone morphogenetic protein 4 | BMP-4 | P12644 | Neuro | 99.17% |
| Eosinophil cationic protein | RSE3 | P12724 | Met | 99.90% |
| Cadherin-1 | CDH1 | P12830 | C-Met | 100.00% |
| Interleukin-7 | IL-7 | P13232 | Inf | 97.93% |
| C-C motif chemokine 4 | CCL4 | P13236 | Inf | 100.00% |
| Monocyte chemotactic protein 1 | MCP-1 | P13500 | Inf, CVD III | 100%; 100% |
| Neural cell adhesion molecule 1 | NCAM1 | P13591 | C-Met | 100.00% |
| Intercellular adhesion molecule 2 | ICAM-2 | P13598 | CVD III | 100.00% |
| Versican core protein | VCAN | P13611 | Met | 100.00% |
| Tartrate-resistant acid phosphatase type 5 | TR-AP | P13686 | CVD III | 100.00% |
| Carcinoembryonic antigen-related cell adhesion molecule 1 | CEACAM1 | P13688 | Onc II | 99.79% |
| Tissue factor | TF | P13726 | CVD II | 100.00% |
| CD59 glycoprotein | CD59 | P13987 | C-Met | 100.00% |
| L-selectin | SELL | P14151 | C-Met | 100.00% |
| Macrophage migration inhibitory factor | MIF | P14174 | Dev | 100.00% |
| Hepatocyte growth factor | HGF | P14210 | Inf, Onc II | 100%; 100% |
| Carboxypeptidase M | CPM | P14384 | Neuro | 99.90% |
| Nidogen-1 | NID1 | P14543 | C-Met | 100.00% |
| Interleukin-1 receptor type 1 | IL-1RT1 | P14778 | CVD III | 100.00% |
| Carboxypeptidase A1 | CPA1 | P15085 | CVD III | 100.00% |
| Carboxypeptidase B | CPB1 | P15086 | CVD III | 100.00% |
| Fatty acid-binding protein, adipocyte (FABP4) | FABP4 | P15090 | CVD III | 100.00% |
| Aminopeptidase N | AP-N | P15144 | CVD III | 100.00% |
| Poliovirus receptor | PVR | P15151 | Neuro | 99.90% |
| Interferon gamma receptor 1 | IFN-gamma-R1 | P15260 | Onc II | 100.00% |
| Arylsulfatase A | ARSA | P15289 | Dev | 100.00% |
| Beta-1,4-galactosyltransferase 1 | B4GALT1 | P15291 | Dev | 100.00% |
| Ezrin | EZR | P15311 | Neuro | 100.00% |
| Folate receptor alpha | FR-alpha | P15328 | Onc II | 100.00% |
| Granulocyte-macrophage colony-stimulating factor receptor subunit alpha | GM-CSF-R-alpha | P15509 | Neuro | 100.00% |
| Amphiregulin | AREG | P15514 | Onc II, IR | 97.42% |
| Membrane cofactor protein | CD46 | P15529 | C-Met | 100.00% |
| Vascular endothelial growth factor A | VEGF-A | P15692 | Inf, Onc II | 100%; 100% |
| Arylsulfatase B | ARSB | P15848 | Cell-Reg | 100.00% |
| Beta-galactoside alpha-2,6-sialyltransferase 1 | ST6GAL1 | P15907 | C-Met | 100.00% |
| P-selectin | SELP | P16109 | CVD III | 100.00% |
| Aggrecan core protein | ACAN | P16112 | Dev | 100.00% |
| Platelet-derived growth factor receptor alpha | PDGF-R-alpha | P16234 | Neuro | 100.00% |
| Beta-galactosidase | GLB1 | P16278 | IR | 97.31% |
| Platelet endothelial cell adhesion molecule | PECAM-1 | P16284 | CVD III | 100.00% |
| Epithelial cell adhesion molecule | Ep-CAM | P16422 | CVD III | 100.00% |
| Cysteine-rich secretory protein 2 | CRISP2 | P16562 | Cell-Reg | 100.00% |
| E-selectin | SELE | P16581 | CVD III | 100.00% |
| Carboxypeptidase E | CPE | P16870 | Onc II | 100.00% |
| Sphingomyelin phosphodiesterase | SMPD1 | P17405 | Neuro | 99.90% |
| Endoglin | ENG | P17813 | C-Met | 95.45% |
| Galectin-3 | Gal-3 | P17931 | CVD III | 100.00% |
| Insulin-like growth factor-binding protein 3 | IGFBP3 | P17936 | C-Met | 100.00% |
| Insulin-like growth factor-binding protein 2 | IGFBP-2 | P18065 | CVD III | 100.00% |
| Integrin beta-5 | ITGB5 | P18084 | Onc II | 100.00% |
| Interleukin-1 receptor antagonist protein | IL-1ra | P18510 | CVD II | 100.00% |
| Syndecan-1 | SYND1 | P18827 | Onc II | 100.00% |
| Peptidyl-glycine alpha-amidating monooxygenase | PAM | P19021 | C-Met | 100.00% |
| Cadherin-2 | CDH2 | P19022 | Met | 100.00% |
| Lymphocyte function-associated antigen 3 | CD58 | P19256 | Dev | 100.00% |
| Vascular cell adhesion protein 1 | VCAM1 | P19320 | C-Met | 100.00% |
| Tumor necrosis factor receptor 1 | TNF-R1 | P19438 | CVD III | 100.00% |
| Follistatin | FS | P19883 | CVD II | 100.00% |
| Elafin | PI3 | P19957 | CVD III | 98.35% |
| Thymidine phosphorylase | TYMP | P19971 | Met | 100.00% |
| Complement receptor type 2 | CR2 | P20023 | C-Met | 97.62% |
| Transcobalamin-2 | TCN2 | P20062 | C-Met | 100.00% |
| Tumor necrosis factor receptor 2 | TNF-R2 | P20333 | CVD III | 100.00% |
| Parvalbumin alpha | PVALB | P20472 | OD | 100.00% |
| Aromatic-L-amino-acid decarboxylase | DDC | P20711 | Met | 99.90% |
| Pleiotrophin | PTN | P21246 | OD | 100.00% |
| Stem cell factor | SCF | P21583 | Inf, CVD II,Onc II | 100%; 100%; 100% |
| 5'-nucleotidase | 5'-NT | P21589 | Onc II | 100.00% |
| Midkine | MK | P21741 | Onc II | 100.00% |
| Macrophage scavenger receptor types I and II | MSR1 | P21757 | Neuro | 99.90% |
| Biglycan | BGN | P21810 | Cell-Reg | 90.49% |
| Receptor tyrosine-protein kinase erbB-3 | ErbB3/HER3 | P21860 | Onc II | 100.00% |
| Protein-glutamine gamma-glutamyltransferase 2 | TGM2 | P21980 | CVD II | 100.00% |
| Bone morphogenetic protein 6 | BMP-6 | P22004 | CVD II | 100.00% |
| Tenascin-X | TNXB | P22105 | C-Met | 94.01% |
| Cadherin-3 | CDH3 | P22223 | Neuro | 99.90% |
| Galanin peptides | GAL | P22466 | Met | 100.00% |
| Carbonic anhydrase 4 | CA4 | P22748 | C-Met | 100.00% |
| Peptidyl-prolyl cis-trans isomerase B | PPIB | P23284 | Dev | 100.00% |
| Oligodendrocyte-myelin glycoprotein | OMG | P23515 | Cell-Reg | 100.00% |
| C-type natriuretic peptide | NPPC | P23582 | OD | 99.90% |
| Corticotropin-releasing factor-binding protein | CRHBP | P24387 | Dev | 99.90% |
| Insulin-like growth factor-binding protein 6 | IGFBP6 | P24592 | C-Met | 100.00% |
| Tenascin | TNC | P24821 | C-Met | 94.32% |
| Proteinase-activated receptor 1 | PAR-1 | P25116 | CVD II | 99.79% |

| | | | | |
|---|---|---|---|---|
| Tumor necrosis factor receptor superfamily member 6 | FAS | P25445 | CVD III | 100.00% |
| Cathepsin S | CTSS | P25774 | Neuro | 100.00% |
| CD40L receptor | CD40 | P25942 | Inf | 100.00% |
| Protein S100-A4 | S100A4 | P26447 | Onc II | 90.39% |
| CD27 antigen | CD27 | P26842 | Onc II | 100.00% |
| Dipeptidyl peptidase 4 | DPP4 | P27487 | C-Met | 99.79% |
| DNA-(apurinic or apyrimidinic site) lyase | APEX1 | P27695 | Met | 93.60% |
| Calreticulin | CALR | P27797 | OD | 85.74% |
| Interleukin-1 receptor type 2 | IL-1RT2 | P27930 | CVD III | 98.45% |
| Cystatin D | CST5 | P28325 | Inf | 100.00% |
| Granulins | GRN | P28799 | CVD III | 100.00% |
| ADP-ribosyl cyclase/cyclic ADP-ribose hydrolase 1 | CD38 | P28907 | Neuro | 100.00% |
| T-cell surface glycoprotein CD1c | CD1C | P29017 | Met | 99.48% |
| Ephrin type-A receptor 2 | EPHA2 | P29317 | Onc II | 100.00% |
| Interleukin-12 subunit beta | IL-12B | P29460 | Inf | 100.00% |
| Interleukin-12 | IL-12 | P29460,P29459 | Neuro | 99.90% |
| Peroxiredoxin-5 | PRDX5 | P30044 | IR | 85.63% |
| Phosphatidylethanolamine-binding protein 1 | PEBP1 | P30086 | Dev | 100.00% |
| Tyrosine-protein kinase receptor UFO | AXL | P30530 | CVD III | 100.00% |
| Alpha-2-macroglobulin receptor-associated protein | Alpha-2-MRAP | P30533 | Neuro | 99.90% |
| Syndecan-4 | SDC4 | P31431 | Met | 100.00% |
| Protein S100-A11 | S100A11 | P31949 | Onc II | 96.90% |
| Glypican-1 | GPC1 | P35052 | Onc II | 100.00% |
| Serpin B6 | SERPINB6 | P35237 | Met | 100.00% |
| ADM | ADM | P35318 | CVD II | 100.00% |
| Thrombospondin-2 | THBS2 | P35442 | CVD II | 100.00% |
| Thrombospondin-4 | THBS4 | P35443 | C-Met | 100.00% |
| Alpha-L-iduronidase | IDUA | P35475 | CVD II | 100.00% |
| Tyrosine-protein kinase receptor Tie-1 | TIE1 | P35590 | C-Met | 100.00% |
| Glutaredoxin-1 | GLRX | P35754 | Met | 100.00% |
| Vascular endothelial growth factor receptor 3 | VEGFR-3 | P35916 | Onc II | 86.98% |
| Vascular endothelial growth factor receptor 2 | VEGFR-2 | P35968 | Onc II | 100.00% |
| Chitinase-3-like protein 1 | CHI3L1 | P36222 | CVD III | 100.00% |
| Lymphotoxin-beta receptor | LTBR | P36941 | CVD III | 100.00% |
| Serine/threonine-protein kinase receptor R3 | SKR3 | P37023 | Neuro | 99.90% |
| TGF-beta receptor type-2 | TGFR-2 | P37173 | Onc II | 100.00% |
| Collagen alpha-1(XVIII) chain | COL18A1 | P39060 | C-Met | 100.00% |
| Matrix metalloproteinase-12 | MMP-12 | P39900 | CVD II | 95.25% |
| Macrophage-capping protein | CAPG | P40121 | OD | 98.76% |
| Alpha-taxilin | TXL | P40222 | Onc II | 99.38% |
| B-cell antigen receptor complex-associated protein beta chain | CD79B | P40259 | Met | 99.17% |
| OX-2 membrane glycoprotein | CD200 | P41217 | Neuro | 100.00% |
| Protein phosphatase inhibitor 2 | PPP1R2 | P41236 | Met | 100.00% |
| Neuroblastoma suppressor of tumorigenicity 1 | NBL1 | P41271 | Neuro | 100.00% |
| Folate receptor gamma | FR-gamma | P41439 | Onc II | 100.00% |
| Caspase-3 | CASP-3 | P42574 | CVD III | 99.90% |
| Dipeptidyl aminopeptidase-like protein 6 | DPP6 | P42658 | OD | 99.90% |
| Leukemia inhibitory factor receptor | LIF-R | P42702 | Inf | 100.00% |
| Lysosomal Pro-X carboxypeptidase | PRCP | P42785 | C-Met | 98.66% |
| C-X-C motif chemokine 5 | CXCL5 | P42830 | Inf | 100.00% |
| Cathepsin O | CTSO | P43234 | Met | 100.00% |
| Tumor necrosis factor receptor superfamily member 4 | TNFRSF4 | P43489 | Onc II | 99.79% |
| Neurogenic locus notch homolog protein 1 | NOTCH1 | P46531 | C-Met | 100.00% |
| Lymphotactin | XCL1 | P47992 | CVD II | 99.90% |
| Fas antigen ligand | FasL | P48023 | Onc II | 100.00% |
| Carboxypeptidase A2 | CPA2 | P48052 | Neuro | 99.90% |
| Tissue factor pathway inhibitor 2 | TFPI-2 | P48307 | Onc II | 100.00% |
| Protein NOV homolog | NOV | P48745 | Dev | 100.00% |
| CD97 antigen | CD97 | P48960 | Dev | 100.00% |
| Cartilage oligomeric matrix protein | COMP | P49747 | C-Met | 100.00% |
| Placenta growth factor | PGF | P49763 | CVD II, OD | 100.00% |
| Fms-related tyrosine kinase 3 ligand | Flt3L | P49771 | Inf | 100.00% |
| Serpin B8 | SERPINB8 | P50452 | Met | 100.00% |
| Methionine aminopeptidase 2 | MetAP 2 | P50579 | Onc II | 98.04% |
| TNF-related apoptosis-inducing ligand | TRAIL | P50591 | Inf, Onc II | 100%; 99,9% |
| Basal cell adhesion molecule | BCAM | P50895 | Dev | 100.00% |
| Eotaxin | CCL11 | P51671 | Inf, IR | 100%; 99,79% |
| Amyloid-like protein 1 | APLP1 | P51693 | Met | 100.00% |
| Prolargin | PRELP | P51888 | CVD II | 100.00% |
| Dual specificity mitogen-activated protein kinase kinase 6 | MAP2K6 | P52564 | Cell-Reg | 96.90% |
| Ephrin-A4 | EF4 | P52798 | Neuro | 99.90% |
| Stanniocalcin-1 | STC1 | P52823 | IR | 100.00% |
| Thimet oligopeptidase | THOP1 | P52888 | Met | 100.00% |
| Dipeptidyl peptidase 1 | CTSC | P53634 | Neuro | 99.90% |
| Ephrin type-B receptor 4 | EPHB4 | P54760 | CVD III | 100.00% |
| Phospholipid transfer protein | PLTP | P55058 | C-Met | 100.00% |
| Mesencephalic astrocyte-derived neurotrophic factor | MANF | P55145 | Neuro | 99.90% |
| Cadherin-6 | CDH6 | P55285 | Neuro | 100.00% |
| C-C motif chemokine 23 | CCL23 | P55773 | Inf | 100.00% |
| C-C motif chemokine 18 | CCL18 | P55774 | C-Met | 100.00% |
| Glycoprotein Xg | XG | P55808 | Dev | 99.79% |
| GDNF family receptor alpha-1 | GFR-alpha-1 | P56159 | Neuro | 99.90% |
| Galectin-4 | Gal-4 | P56470 | CVD III | 90.29% |
| Junctional adhesion molecule B | JAM-B | P57087 | Neuro | 100.00% |
| Protein FAM3B | FAM3B | P58499 | IR | 100.00% |
| Thymosin beta-10 | TMSB10 | P63313 | Dev | 100.00% |
| Coxsackievirus and adenovirus receptor | CXADR | P78310 | IR | 100.00% |
| Tyrosine-protein phosphatase non-receptor type substrate 1 | SHPS-1 | P78324 | CVD III | 99.90% |
| Disintegrin and metalloproteinase domain-containing protein 8 | ADAM8 | P78325 | Onc II | 99.28% |
| Glypican-5 | GCP5 | P78333 | Neuro | 100.00% |
| Lectin-like oxidized LDL receptor 1 | LOX-1 | P78380 | CVD II | 100.00% |
| Butyrophilin subfamily 3 member A2 | BTN3A2 | P78410 | IR | 100.00% |
| Fractalkine | CX3CL1 | P78423 | Inf | 100.00% |
| Interleukin-13 receptor subunit alpha-1 | IL13RA1 | P78552 | Dev | 94.11% |
| Monocyte chemotactic protein 2 | MCP-2 | P80075 | Inf | 100.00% |
| C-X-C motif chemokine 6 | CXCL6 | P80162 | Inf | 100.00% |
| Nucleobindin-2 | NUCB2 | P80303 | OD | 99.79% |
| Protein delta homolog 1 | DLK-1 | P80370 | CVD III | 100.00% |
| Perlecan | PLC | P98160 | CVD III | 100.00% |
| CD83 antigen | CD83 | Q01151 | IR | 100.00% |
| ST2 protein | ST2 | Q01638 | CVD III | 93.70% |
| Inactive tyrosine-protein kinase transmembrane receptor ROR1 | ROR1 | Q01973 | Met | 100.00% |
| N-acylethanolamine-hydrolyzing acid amidase | NAAA | Q02083 | Neuro | 99.69% |
| Contactin-2 | CNTN2 | Q02246 | OD | 100.00% |
| Desmocollin-2 | DSC2 | Q02487 | Dev | 100.00% |
| Angiopoietin-1 receptor | TIE2 | Q02763 | CVD II | 100.00% |
| Peptidyl-prolyl cis-trans isomerase FKBP4 | FKBP4 | Q02790 | Met | 100.00% |
| Transforming growth factor beta receptor type 3 | TGFBR3 | Q03167 | C-Met | 100.00% |
| Trefoil factor 2 | TFF2 | Q03403 | Met | 99.90% |
| Urokinase plasminogen activator surface receptor | U-PAR | Q03405 | CVD III | 100.00% |
| Parathyroid hormone/parathyroid hormone-related peptide receptor | PTH1R | Q03431 | IR | 86.04% |
| Lactoylglutathione lyase | GLO1 | Q04760 | CVD II | 100.00% |
| Sialomucin core protein 24 | CD164 | Q04900 | Met | 100.00% |
| Tyrosine-protein kinase receptor TYRO3 | TYRO3 | Q06418 | Met | 100.00% |
| Peroxiredoxin-1 | PRDX1 | Q06830 | IR | 100.00% |
| Tumor necrosis factor receptor superfamily member 9 | TNFRSF9 | Q07011 | Inf | 100.00% |
| Cytoskeleton-associated protein 4 | CKAP4 | Q07065 | IR | 100.00% |
| Early activation antigen CD69 | CD69 | Q07108 | Dev | 100.00% |
| C-X-C motif chemokine 9 | CXCL9 | Q07325 | Inf | 100.00% |
| Trefoil factor 3 | TFF3 | Q07654 | CVD III | 100.00% |
| Interleukin-10 receptor subunit beta | IL-10RB | Q08334 | Inf | 100.00% |
| Epithelial discoidin domain-containing receptor 1 | DDR1 | Q08345 | Neuro | 100.00% |
| Lactadherin | MFGE8 | Q08431 | Dev | 100.00% |
| Testican-1 | SPOCK1 | Q08629 | Neuro | 100.00% |
| CMRF35-like molecule 6 | CLM-6 | Q08708 | Neuro | 99.90% |
| Polypeptide N-acetylgalactosaminyltransferase 2 | GALNT2 | Q10471 | Cell-Reg | 100.00% |
| CMP-N-acetylneuraminate-beta-galactosamide-alpha-2,3-sialyltransferase 1 | ST3GAL1 | Q11201 | OD | 99.69% |
| EGF-containing fibulin-like extracellular matrix protein 1 | EFEMP1 | Q12805 | C-Met | 100.00% |
| Contactin-1 | CNTN1 | Q12860 | CVD III | 100.00% |
| Tyrosine-protein kinase Mer | MERTK | Q12866 | CVD II | 95.76% |
| BMP and activin membrane-bound inhibitor homolog | BAMBI | Q13145 | OD | 99.79% |
| Pappalysin-1 | PAPPA | Q13219 | CVD II | 100.00% |
| Chitotriosidase-1 | CHIT1 | Q13231 | CVD III | 96.38% |
| Natural killer cells antigen CD94 | KLRD1 | Q13241 | IR | 92.55% |
| Semaphorin-3F | SEMA3F | Q13275 | Met | 100.00% |
| Inactive tyrosine-protein kinase 7 | PTK7 | Q13308 | OD | 94.52% |
| Receptor-type tyrosine-protein phosphatase S | PTPRS | Q13332 | C-Met | 100.00% |
| Microfibrillar-associated protein 5 | MFAP5 | Q13361 | C-Met | 99.79% |

| | | | | |
|---|---|---|---|---|
| Mesothelin | MSLN | Q13421 | Onc II | 99.90% |
| Interleukin-18 receptor 1 | IL-18R1 | Q13478 | Inf | 100.00% |
| Eukaryotic translation initiation factor 4E-binding protein 1 | 4E-BP1 | Q13541 | Inf | 100.00% |
| CD166 antigen | ALCAM | Q13740 | CVD III | 100.00% |
| Bleomycin hydrolase | BLM hydrolase | Q13867 | CVD III | 100.00% |
| Pro-interleukin-16 | IL16 | Q14005 | CVD II | 94.32% |
| Lysosome membrane protein 2 | SCARB2 | Q14108 | Neuro | 99.90% |
| Nidogen-2 | NID2 | Q14112 | Dev | 100.00% |
| Interleukin-18 | IL-18 | Q14116 | Inf, CVD II | 100%; 99,59% |
| Dystroglycan | DAG1 | Q14118 | Dev | 100.00% |
| Scavenger receptor class F member 1 | SCARF1 | Q14162 | Dev | 100.00% |
| P-selectin glycoprotein ligand 1 | PSGL-1 | Q14242 | CVD II | 100.00% |
| Growth arrest-specific protein 6 | GAS6 | Q14393 | C-Met | 100.00% |
| WAP four-disulfide core domain protein 2 | WFDC2 | Q14508 | Onc II | 100.00% |
| SPARC-like protein 1 | SPARCL1 | Q14515 | C-Met | 100.00% |
| LDLR chaperone MESD | MESDC2 | Q14696 | Dev | 100.00% |
| Latent-transforming growth factor beta-binding protein 2 | LTBP2 | Q14767 | C-Met | 100.00% |
| Procollagen C-endopeptidase enhancer 1 | PCOLCE | Q15113 | C-Met | 100.00% |
| Nodal modulator 1 | NOMO1 | Q15155 | Met | 100.00% |
| Serum paraoxonase/arylesterase 2 | PON2 | Q15165 | OD | 94.94% |
| Paraoxonase | PON3 | Q15166 | CVD III | 92.87% |
| Receptor tyrosine-protein kinase erbB-4 | ErbB4/HER4 | Q15303 | Onc II | 100.00% |
| Angiopoietin-1 | ANG-1 | Q15389 | CVD II | 100.00% |
| Ficolin-2 | FCN2 | Q15485 | C-Met | 92.36% |
| Transforming growth factor-beta-induced protein ig-h3 | TGFBI | Q15582 | C-Met | 100.00% |
| Cystatin-M | CST6 | Q15828 | Dev | 100.00% |
| Clusterin-like protein 1 | CLUL1 | Q15846 | Met | 100.00% |
| Insulin-like growth factor-binding protein 7 | IGFBP-7 | Q16270 | CVD III | 100.00% |
| NT-3 growth factor receptor | NTRK3 | Q16288 | Neuro | 99.90% |
| Laminin subunit alpha-4 | LAMA4 | Q16363 | Dev | 100.00% |
| BDNF/NT-3 growth factors receptor | NTRK2 | Q16620 | Neuro | 100.00% |
| C-C motif chemokine 14 | CCL14 | Q16627 | C-Met | 100.00% |
| Prostasin | PRSS8 | Q16651 | CVD II | 100.00% |
| Myelin-oligodendrocyte glycoprotein | MOG | Q16653 | Cell-Reg | 100.00% |
| C-C motif chemokine 15 | CCL15 | Q16663 | CVD III | 100.00% |
| Melanoma-derived growth regulatory protein | MIA | Q16674 | Onc II | 100.00% |
| Kynureninase | KYNU | Q16719 | Neuro | 100.00% |
| Glutaminyl-peptide cyclotransferase | QPCT | Q16769 | C-Met | 100.00% |
| Kynurenine--oxoglutarate transaminase 1 | KYAT1 | Q16773 | Met | 100.00% |
| Carbonic anhydrase IX | CAIX | Q16790 | Onc II | 100.00% |
| Membrane primary amine oxidase | AOC3 | Q16853 | C-Met | 99.59% |
| MHC class I polypeptide-related sequence A/B | MIC-A/B | Q29983,Q29980 | Onc II | 99.59% |
| R-spondin-1 | RSPO1 | Q2MKA7 | Neuro | 98.76% |
| Brorin | VWC2 | Q2TAL6 | Neuro | 99.90% |
| Protogenin | PRTG | Q2VWP7 | Neuro | 99.90% |
| Cell adhesion molecule-related/down-regulated by oncogenes | CDON | Q4KMG0 | Dev | 99.79% |
| Collectin-12 | COLEC12 | Q5KU26 | Dev | 100.00% |
| Platelet endothelial aggregation receptor 1 | PEAR1 | Q5VY43 | Dev | 100.00% |
| Meteorin-like protein | METRNL | Q641Q3 | Met | 100.00% |
| Vasorin | VASN | Q6EMK4 | C-Met | 100.00% |
| Leukocyte-associated immunoglobulin-like receptor 1 | LAIR1 | Q6GTX8 | Dev | 100.00% |
| Leukocyte-associated immunoglobulin-like receptor 2 | LAIR-2 | Q6ISS4 | Neuro | 99.48% |
| RGM domain family member B | RGMB | Q6NW40 | Neuro | 100.00% |
| Layilin | LAYN | Q6UX15 | Neuro | 100.00% |
| VEGF-co regulated chemokine 1 | CXL17 | Q6UXB2 | Onc II | 99.28% |
| Seizure 6-like protein 2 | SEZ6L2 | Q6UXD5 | Cell-Reg | 100.00% |
| CMRF35-like molecule 9 | CD300LG | Q6UXG3 | Dev | 97.31% |
| Cysteine-rich with EGF-like domain protein 2 | CRELD2 | Q6UXH1 | Dev | 100.00% |
| Inactive serine protease PAMR1 | PAMR1 | Q6UXH9 | Dev | 100.00% |
| Leucine-rich repeat neuronal protein 1 | LRRN1 | Q6UXK5 | Cell-Reg | 100.00% |
| Chordin-like protein 2 | CHRDL2 | Q6WN34 | Met | 95.56% |
| CD109 antigen | CD109 | Q6YHK3 | Dev | 100.00% |
| Scavenger receptor class A member 5 | SCARA5 | Q6ZMJ2 | Neuro | 99.90% |
| Coiled-coil domain-containing protein 80 | CCDC80 | Q76M96 | Met | 100.00% |
| Protein FAM19A5 | FAM19A5 | Q7Z5A7 | Cell-Reg | 100.00% |
| Allergin-1 | MILR1 | Q7Z6M3 | IR | 100.00% |
| Amphoterin-induced protein 2 | AMIGO2 | Q86SJ2 | Cell-Reg | 100.00% |
| Polypeptide N-acetylgalactosaminyltransferase 10 | GALNT10 | Q86SR1 | OD | 99.79% |
| C-type lectin domain family 14 member A | CLEC14A | Q86T13 | Dev | 100.00% |
| Scavenger receptor cysteine-rich type 1 protein M130 | CD163 | Q86VB7 | CVD III | 100.00% |
| Low-density lipoprotein receptor-related protein 11 | LRP11 | Q86VZ4 | Met | 100.00% |
| Interferon lambda receptor 1 | IFNLR1 | Q8IU57 | IR | 99.69% |
| Plexin domain-containing protein 1 | PLXDC1 | Q8IUK5 | OD | 99.79% |
| Contactin-4 | CNTN4 | Q8IWV2 | Dev | 100.00% |
| SIR2-like protein 2 | SIRT2 | Q8IXJ6 | Inf | 97.73% |
| Osteoclast-associated immunoglobulin-like receptor | hOSCAR | Q8IYS5 | CVD II | 100.00% |
| Cell adhesion molecule 3 | CADM3 | Q8N126 | Neuro | 100.00% |
| Leukocyte immunoglobulin-like receptor subfamily B member 2 | LILRB2 | Q8N423 | C-Met | 99.59% |
| Inactive dipeptidyl peptidase 10 | DPP10 | Q8N608 | IR | 100.00% |
| CD177 antigen | CD177 | Q8N6Q3 | Dev | 95.66% |
| Draxin | DRAXIN | Q8NBI3 | Neuro | 100.00% |
| Sulfatase-modifying factor 2 | SUMF2 | Q8NBJ7 | Met | 100.00% |
| Proprotein convertase subtilisin/kexin type 9 | PCSK9 | Q8NBP7 | CVD III | 99.79% |
| Thioredoxin domain-containing protein 5 | TXNDC5 | Q8NBS9 | Met | 100.00% |
| Interleukin-27 | IL-27 | Q8NEV9,Q14213 | CVD II | 100.00% |
| MAM domain-containing glycosylphosphatidylinositol anchor protein 1 | MDGA1 | Q8NFP4 | Neuro | 100.00% |
| Leukocyte immunoglobulin-like receptor subfamily B member 4 | LILRB4 | Q8NHJ6 | IR | 100.00% |
| Leukocyte immunoglobulin-like receptor subfamily B member 1 | LILRB1 | Q8NHL6 | C-Met | 100.00% |
| Multiple coagulation factor deficiency protein 2 | MCFD2 | Q8NI22 | Met | 100.00% |
| Interleukin-17D | IL-17D | Q8TAD2 | CVD II | 100.00% |
| CD99 antigen-like protein 2 | CD99L2 | Q8TCZ2 | Dev | 100.00% |
| Cell surface glycoprotein CD200 receptor 1 | CD200R1 | Q8TD46 | Neuro | 98.04% |
| Hepatitis A virus cellular receptor 2 | HAVCR2 | Q8TDQ0 | Dev | 100.00% |
| CMRF35-like molecule 1 | CLM-1 | Q8TDQ1 | Neuro | 99.90% |
| A disintegrin and metalloproteinase with thrombospondin motifs 15 | ADAM-TS 15 | Q8TE58 | Onc II | 100.00% |
| WAP, Kazal, immunoglobulin, Kunitz and NTR domain-containing protein 2 | WFIKKN2 | Q8TEU8 | Dev | 100.00% |
| Soluble calcium-activated nucleotidase 1 | CANT1 | Q8WVQ1 | Met | 100.00% |
| Insulin-like growth factor-binding protein-like 1 | IGFBPL1 | Q8WX77 | Met | 100.00% |
| Protein FAM3C | FAM3C | Q92520 | Met | 100.00% |
| C-C motif chemokine 17 | CCL17 | Q92583 | CVD II | 99.90% |
| Nectin-2 | NECTIN2 | Q92692 | Met | 100.00% |
| Tenascin-R | TN-R | Q92752 | Neuro | 100.00% |
| Secreted frizzled-related protein 3 | sFRP-3 | Q92765 | Neuro | 100.00% |
| Neuronal cell adhesion molecule | Nr-CAM | Q92823 | Neuro | 100.00% |
| Kallikrein-6 | KLK6 | Q92876 | CVD III | 100.00% |
| Tumor necrosis factor receptor superfamily member 14 | TNFRSF14 | Q92956 | CVD III | 100.00% |
| Tumor necrosis factor receptor superfamily member 19L | RELT | Q969Z4 | Dev | 100.00% |
| Endothelial cell-selective adhesion molecule | ESAM | Q96AP7 | Dev | 100.00% |
| Repulsive guidance molecule A | RGMA | Q96B86 | Neuro | 100.00% |
| Kidney Injury Molecule | KIM1 | Q96D42 | CVD II, OD | 100%; 97,12% |
| Interleukin-17 receptor A | IL-17RA | Q96F46 | CVD III | 100.00% |
| Scavenger receptor class F member 2 | SCARF2 | Q96GP6 | Neuro | 99.90% |
| Brevican core protein | BCAN | Q96GW7 | Neuro | 100.00% |
| T-cell immunoglobulin and mucin domain-containing protein 4 | TIMD4 | Q96H15 | C-Met | 91.01% |
| Kazal-type serine protease inhibitor domain-containing protein 1 | KAZALD1 | Q96I82 | Cell-Reg | 99.59% |
| Leucine-rich repeats and immunoglobulin-like domains protein 1 | LRIG1 | Q96JA1 | Met | 100.00% |
| Beta-Ala-His dipeptidase | CNDP1 | Q96KN2 | C-Met | 100.00% |
| Fc receptor-like protein 1 | FCRL1 | Q96LA6 | Met | 92.77% |
| Sialic acid-binding Ig-like lectin 10 | SIGLEC10 | Q96LC7 | Cell-Reg | 100.00% |
| Nectin-4 | PVRL4 | Q96NY8 | Onc II | 99.90% |
| WAP, Kazal, immunoglobulin, Kunitz and NTR domain-containing protein 1 | WFIKKN1 | Q96NZ8 | Neuro | 92.25% |
| Discoidin, CUB and LCCL domain-containing protein 2 | DCBLD2 | Q96PD2 | IR | 93.38% |
| Secretoglobin family 3A member 2 | SCGB3A2 | Q96PL1 | CVD III | 86.98% |
| VPS10 domain-containing receptor SorCS2 | SORCS2 | Q96PQ0 | Cell-Reg | 100.00% |
| Probable carboxypeptidase X1 | CPXM1 | Q96SM3 | Cell-Reg | 100.00% |
| Proheparin-binding EGF-like growth factor | HB-EGF | Q99075 | CVD II | 100.00% |
| Protein deglycase DJ-1 | PARK7 | Q99497 | Dev | 100.00% |
| Sortilin | SORT1 | Q99523 | CVD II | 100.00% |
| Legumain | LGMN | Q99538 | Dev | 100.00% |
| Oncostatin-M-specific receptor subunit beta | OSMR | Q99650 | C-Met | 99.90% |
| Mothers against decapentaplegic homolog 5 | MAD homolog 5 | Q99717 | Onc II | 100.00% |
| Metalloproteinase inhibitor 4 | TIMP4 | Q99727 | CVD III | 100.00% |
| C-C motif chemokine 19 | CCL19 | Q99731 | Inf | 100.00% |
| Chymotrypsin C | CTRC | Q99895 | CVD II | 98.86% |
| Retinoic acid receptor responder protein 2 | RARRES2 | Q99969 | CVD III | 100.00% |
| Myocilin | MYOC | Q99972 | Dev | 100.00% |
| Osteomodulin | OMD | Q99983 | Dev | 100.00% |
| Growth/differentiation factor 15 | GDF-15 | Q99988 | CVD III | 100.00% |

| | | | | |
|---|---|---|---|---|
| Sclerostin | SOST | Q9BQB4 | Met | 100.00% |
| Serine protease 27 | PRSS27 | Q9BQR3 | CVD II | 100.00% |
| Calsyntenin-3 | CLSTN3 | Q9BQT9 | Cell-Reg | 100.00% |
| Spondin-2 | SPON2 | Q9BUD6 | CVD II | 100.00% |
| Brother of CDO | Protein BOC | Q9BWV1 | CVD II, Cell-Reg | 100%; 100% |
| Complement C1q tumor necrosis factor-related protein 1 | C1QTNF1 | Q9BXJ1 | C-Met | 100.00% |
| C-type lectin domain family 7 member A | CLEC7A | Q9BXN2 | IR | 99.28% |
| Complement factor H-related protein 5 | CFHR5 | Q9BXR6 | C-Met | 100.00% |
| R-spondin-3 | RSPO3 | Q9BXY4 | Onc II | 100.00% |
| Angiopoietin-related protein 4 | ANGPTL4 | Q9BY76 | Dev | 100.00% |
| Seizure 6-like protein | SEZ6L | Q9BYH1 | Onc II | 100.00% |
| Regenerating islet-derived protein 4 | REG4 | Q9BYZ8 | Met | 100.00% |
| NKG2D ligand 2 | N2DL-2 | Q9BZM5 | Neuro | 99.90% |
| Reticulon-4 receptor | RTN4R | Q9BZR6 | Met | 100.00% |
| Natural killer cell receptor 2B4 | CD244 | Q9BZW8 | Inf | 99.48% |
| Sialoadhesin | SIGLEC1 | Q9BZZ2 | Neuro | 99.90% |
| Semaphorin-4C | SEMA4C | Q9C0C4 | Cell-Reg | 99.90% |
| Tubulointerstitial nephritis antigen-like | TIGL1 | Q9GZM7 | Met | 100.00% |
| SLIT and NTRK-like protein 2 | SLITRK2 | Q9H156 | Cell-Reg | 100.00% |
| Multiple epidermal growth factor-like domains protein 9 | MEGF9 | Q9H1U4 | C-Met | 100.00% |
| C-X-C motif chemokine 16 | CXCL16 | Q9H2A7 | CVD III | 100.00% |
| Transmembrane protease serine 5 | TMPRSS5 | Q9H3S3 | Neuro | 100.00% |
| SPARC-related modular calcium-binding protein 2 | SMOC2 | Q9H3U7 | Neuro | 100.00% |
| Calsyntenin-2 | CLSTN2 | Q9H4D0 | Met | 100.00% |
| CUB domain-containing protein 1 | CDCP1 | Q9H5V8 | Inf | 100.00% |
| SLIT and NTRK-like protein 6 | SLITRK6 | Q9H5Y7 | Cell-Reg | 91.83% |
| CXADR-like membrane protein | CLMP | Q9H6B4 | Met | 100.00% |
| Tumor necrosis factor receptor superfamily member 27 | EDA2R | Q9HAV5 | Neuro | 99.90% |
| Interleukin-1 receptor-like 2 | IL1RL2 | Q9HB29 | CVD II | 99.79% |
| T-lymphocyte surface antigen Ly-9 | LY9 | Q9HBG7 | Onc II | 89.88% |
| Spondin-1 | SPON1 | Q9HCB6 | CVD III | 100.00% |
| Roundabout homolog 2 | ROBO2 | Q9HCK4 | Neuro | 99.90% |
| Plexin-A4 | PLX4 | Q9HCM2 | IR | 100.00% |
| Resistin | RETN | Q9HD89 | CVD III | 100.00% |
| CD209 antigen | CD209 | Q9NNX6 | Dev | 100.00% |
| Tumor necrosis factor receptor superfamily member 12A | TNFRSF12A | Q9NP84 | Neuro | 99.90% |
| Triggering receptor expressed on myeloid cells 1 | TREM1 | Q9NP99 | IR | 98.76% |
| Lysophosphatidic acid phosphatase type 6 | ACP6 | Q9NPH0 | Met | 100.00% |
| Complement component C1q receptor | CD93 | Q9NPY3 | CVD III | 100.00% |
| Endothelial cell-specific molecule 1 | ESM-1 | Q9NQ30 | Onc II | 100.00% |
| Cartilage acidic protein 1 | CRTAC1 | Q9NQ79 | C-Met | 100.00% |
| Fructose-2,6-bisphosphatase TIGAR (TIGAR) | TIGAR | Q9NQ88 | OD | 88.43% |
| Neural proliferation differentiation and control protein 1 | NPDC1 | Q9NQX5 | Met | 100.00% |
| Neutral ceramidase | N-CDase | Q9NR71 | Neuro | 99.69% |
| C-C motif chemokine 28 | CCL28 | Q9NRJ3 | Inf | 97.00% |
| Interleukin-17 receptor B | IL17RB | Q9NRM6 | Cell-Reg | 100.00% |
| Tumor necrosis factor receptor superfamily member 19 | TNFRSF19 | Q9NS68 | Onc II | 100.00% |
| Fibroblast growth factor 21 | FGF-21 | Q9NSA1 | Cell-Reg | 86.76% |
| Phosphoprotein associated with glycosphingolipid-enriched microdomains 1 | PAG1 | Q9NWQ8 | Met | 100.00% |
| C-type lectin domain family 5 member A | CLEC5A | Q9NY25 | Met | 100.00% |
| Podocalyxin-like protein 2 | PODXL2 | Q9NZ53 | Cell-Reg | 100.00% |
| Programmed cell death 1 ligand 1 | PD-L1 | Q9NZQ7 | Inf | 100.00% |
| Cysteine-rich motor neuron 1 protein | CRIM1 | Q9NZV1 | Dev | 100.00% |
| Kallikrein-14 | hK14 | Q9P0G3 | Onc II | 96.80% |
| Disintegrin and metalloproteinase domain-containing protein 22 | ADAM 22 | Q9P0K1 | Neuro | 100.00% |
| C-type lectin domain family 1 member B | CLEC1B | Q9P126 | Neuro | 97.52% |
| Dickkopf-related protein 3 | DKK3 | Q9UBP4 | Dev | 100.00% |
| Cathepsin Z | CTSZ | Q9UBR2 | CVD III | 100.00% |
| Dickkopf-related protein 4 | Dkk-4 | Q9UBT3 | Neuro | 99.90% |
| Cathepsin F | CTSF | Q9UBX1 | Dev | 100.00% |
| Kallikrein-11 | hK11 | Q9UBX7 | Onc II | 100.00% |
| Macrophage receptor MARCO | MARCO | Q9UEW3 | CVD II | 100.00% |
| Contactin-associated protein-like 2 | CNTNAP2 | Q9UHC6 | IR | 100.00% |
| Epidermal growth factor-like protein 7 | EGFL7 | Q9UHF1 | OD | 99.59% |
| Dipeptidyl peptidase 2 | DPP7 | Q9UHL4 | Met | 99.48% |
| Adhesion G protein-coupled receptor E2 | ADGRE2 | Q9UHX3 | Met | 100.00% |
| SLAM family member 5 | CD84 | Q9UIB8 | CVD II | 99.38% |
| Paired immunoglobulin-like type 2 receptor beta | PILRB | Q9UKJ0 | Met | 100.00% |
| Paired immunoglobulin-like type 2 receptor alpha | PILRA | Q9UKJ1 | Dev | 100.00% |
| ADP-sugar pyrophosphatase | NUDT5 | Q9UKK9 | Dev | 97.93% |
| Kallikrein-13 | KLK13 | Q9UKR3 | Onc II | 100.00% |
| Plexin-B3 | PLXNB3 | Q9ULL4 | Neuro | 100.00% |
| Carbonic anhydrase 14 | CA14 | Q9ULX7 | OD | 99.79% |
| Neurogenic locus notch homolog protein 3 | Notch 3 | Q9UM47 | CVD III | 100.00% |
| Tumor necrosis factor receptor superfamily member EDAR | EDAR | Q9UNE0 | IR | 99.07% |
| Syntaxin-8 | STX8 | Q9UNK0 | OD | 99.79% |
| C-type lectin domain family 11 member A | CLEC11A | Q9Y240 | Dev | 100.00% |
| Tumor necrosis factor ligand superfamily member 13B | TNFSF13B | Q9Y275 | CVD III | 100.00% |
| V-set and immunoglobulin domain-containing protein 4 | VSIG4 | Q9Y279 | Dev | 99.90% |
| Sialic acid-binding Ig-like lectin 7 | SIGLEC7 | Q9Y286 | Met | 100.00% |
| Sialic acid-binding Ig-like lectin 9 | Siglec-9 | Q9Y336 | Neuro | 100.00% |
| CD2-associated protein | CD2AP | Q9Y5K6 | Met | 92.67% |
| Ectonucleoside triphosphate diphosphohydrolase 2 | ENTPD2 | Q9Y5L3 | OD | 99.48% |
| Wnt inhibitory factor 1 | WIF-1 | Q9Y5W5 | Onc II | 100.00% |
| Lymphatic vessel endothelial hyaluronic acid receptor 1 | LYVE1 | Q9Y5Y7 | C-Met | 100.00% |
| Junctional adhesion molecule A | JAM-A | Q9Y624 | CVD III | 100.00% |
| Adhesion G-protein coupled receptor G1 | ADGRG1 | Q9Y653 | OD | 99.38% |
| Roundabout homolog 1 | ROBO1 | Q9Y6N7 | Dev | 100.00% |
| Tumor necrosis factor receptor superfamily member 11A | TNFRSF11A | Q9Y6Q6 | CVD II | 100.00% |
| Delta and Notch-like epidermal growth factor-related receptor | DNER | Q8NFT8 | Inf | 100.00% |
| Transmembrane glycoprotein NMB | GPNMB | Q14956 | Onc II | 100.00% |
| Galactoside 3(4)-L-fucosyltransferase,Alpha-(1,3)-fucosyltransferase 3/5 | (FUT3/5) | Q11128,P21217 | Dev | 95.86% |
| Fibroblast growth factor 5 (FGF-5) | FGF-5 | P12034 | Inf | 100.00% |

Table shows those proteins that passed the quslity control criteria and were detected in more than 85% of the samples. Panel Key: CVD II = Cardiovascular disease II; CVD III =Cardiovasculr disease III; C-Met = Cardiometabolic; Cell-Reg = Cell Regulation; Dev = Development; Inf = Inflammation; IR = Immune Response; I/O = Immuno-Oncology; Met = Metabolism; Neuro = Neurology; OD = Organ Damage; Onc II = Oncology II

**Supplementary Table 3. Quality parameters of custom PEA assays**

| Protein name | Assay | Uniport | LOD | CSF missingness (%) | Intra-CV (%) | Inter-CV (%) |
|---|---|---|---|---|---|---|
| Aromatic-L-amino-acid decarboxylase | DDC | P20711 | 1.98 | 1% | 5% | 8% |
| Corticoliberin | CRH | P06850 | 1.60 | 1% | 7% | 18% |
| Matrix metalloproteinase-3 | MMP-3 | P08254 | 0.38 | 1% | 4% | 8% |
| Tyrosine-protein kinase ABL1 | ABL1* | P00519 | 1.01 | 10% | 5% | 2% |
| Matrix metalloproteinase-10 | MMP-10 | P09238 | 0.98 | 1% | 5% | 8% |
| Thimet oligopeptidase | THOP1 | P52888 | 1.38 | 1% | 5% | 8% |
| | | | Mean | 3% | 5% | 9% |

CSF missingness is the % of samples whose values were < the corresponding LOD. CVs caculated using quality control samples. *For ABL1 CVs were calculated using assay calibrators, as QC samples reported values <LOD. LOD, Lower limit of detection. CV, coefficient of variation.
